# Maternal cigarette smoking and cleft lip and palate: A systematic review and meta-analysis

**DOI:** 10.1101/2021.06.10.21258688

**Authors:** Matthew Fell, Kyle Dack, Shaheel Chummun, Jonathan Sandy, Yvonne Wren, Sarah Lewis

## Abstract

**Objectives:** A systematic review and meta-analysis to determine the association between active maternal smoking and cleft lip and palate etiology.

**Data Sources:** Medline, Embase, Web of Science and the Cochrane database from inception to November 2020.

**Study selection:** Observational studies of cigarette smoking habits in pregnant women. Outcomes included cleft lip and/or palate, cleft lip ± palate and cleft palate only.

**Data analysis:** Publication bias analyses were performed and the Newcastle Ottawa scales were used to assess study quality. Fixed or random effect models were used in the meta-analysis, dependent on risk of statistical heterogeneity.

**Results:** Forty-five studies were eligible for inclusion of which 11 were cohort and 34 were case-control studies. Sixteen studies were of sufficient standard for inclusion in the meta-analysis. The summary odds ratio for the association between smoking and cleft lip and/or palate was 1.42 (95%CI 1.27 to 1.59) with a population attributable fraction of 4% (95%CI 3% - 5%). There was limited evidence to show a dose-response effect of smoking.

**Conclusions:** This review reports a moderate association between maternal smoking and orofacial cleft but the overall quality of the conventional observational studies included was poor. There is a need for high quality and novel research strategies to further define the role of smoking in the etiology of cleft lip and palate.

## INTRODUCTION

Cleft lip and/or palate (CL/P) is one of the most common craniofacial birth defects, occurring in approximately 1/700 births (Mossey et al., 2009). It affects children and their families because of appearance and functional difficulties with speech, eating, social interaction and child development. Seventy percent of children born with CL/P do not have an associated syndrome and the anomaly is believed to be caused by a complex pattern of inheritance with both genetic and environmental influences (Lebby et al., 2010). Defining the role of potentially modifiable environmental factors could reduce the incidence of this congenital abnormality (Raut et al., 2019). Maternal smoking is a modifiable environmental factor, which is considered a causal factor for CL/P in the 2014 US Surgeon General’s Report (United States Department of Health and Human Services 2014).

Cigarette smoke is a complex aerosol comprising more than 4,000 different compounds that can cause harm (Martelli et al., 2015). Maternal smoking has attracted research interest because it is a common exposure and has been established as a risk factor for a spectrum of adverse offspring outcomes including preterm birth, low birth weight and birth anomalies (Krueger and Rohrich 2001; Hackshaw et al., 2011). It is biologically plausible that maternal smoking could cause CL/P, although the exact mechanism is unknown (Leite et al., 2002; Krapels et al., 2008). There may be a direct interaction of the smoking products with neonatal tissue, leading to induced hypoxia because of impaired angiogenesis and nicotine-mediated vasoconstriction, which has been shown to disrupt palatal fusion in animal models (Vieira and Dattilo, 2018). An alternative theory is that smoking affects DNA methylation in the fetus, which could impact upon gene expression responsible for lip and palate formation (Lebby et al., 2010).

Three previous meta-analyses have demonstrated weak to moderate links between maternal smoking and CL/P (Wyszynski et al., 1997; Julian Little et al., 2004; Xuan et al., 2016;). Whilst previous systematic reviews have been comprehensive, the included studies were not assessed for their quality and this might have compromised the validity of the findings (Crossan and Duane, 2018). Potential sources of bias in the primary studies include no adjustment for confounders, inappropriate control groups and recall bias. There is a need for an updated systematic review with rigorous methodology in this field. We conducted a systematic review and meta-analysis in order to determine the role of active maternal cigarette smoking in the etiology of CL/P.

## METHODS

### Identification of studies

A full protocol of this systematic review, carried out following PRISMA guidance(Moher et al., 2009), was adhered to (see supplementary Table 1) and is available from the PROSPERO systematic review register (registration number CRD42020222837; https://www.crd.york.ac.uk/prospero/display_record.php?ID=CRD42020222837).

Eligible studies were defined as full-text primary-data publications reporting on pregnant women from the general population who were assessed for pre-natal active cigarette smoking. Studies were required to document maternal smoking (either in the peri-conception period or any of the three trimesters) but the assessment of smoking status could have been performed prospectively or retrospectively. Studies of passive (or environmental) maternal smoking or paternal smoking were not included. The protocol included all epidemiological studies using an analytical design whereby an exposed group was compared to an unexposed group. Cohort, case-control, quasi-experimental, natural experiment, family based negative control and Mendelian Randomization study designs were eligible.

The outcome of interest was a live born child with CL/P or subtypes such as cleft lip only, cleft lip ± palate (CL±P), cleft palate only (CP) or submucous cleft. Where studies made a distinction between children born with an isolated cleft or a cleft co-occurring with other anomalies, or where results were provided for those with non-syndromic and syndromic orofacial clefts separately, effect estimates for isolated and non-syndromic clefts were extracted preferentially.

Studies were excluded if: full text was unavailable; they were conference proceedings only; they were descriptive studies such as case studies, case series, cross-sectional studies, expert opinion, letter, editorials or studies using secondary data such as reviews; they were animal studies; or there was insufficient data to estimate the effect size of the association between maternal smoking and CL/P (see Supplementary Table 2 for exclusion and exclusion criteria).

The databases searched included Medline, Embase, the Web of Science and the Cochrane Library from inception to 9^th^ November 2020. The search was tailored individually to each database with input from a University Librarian (see Supplementary Figures 1-4 for search strategies) and there was no language restriction. The search focused on published literature and did not include grey literature. In addition, manual searches of reference lists of recent relevant systematic reviews and all studies included in the systematic review were performed.

**Figure 1:**
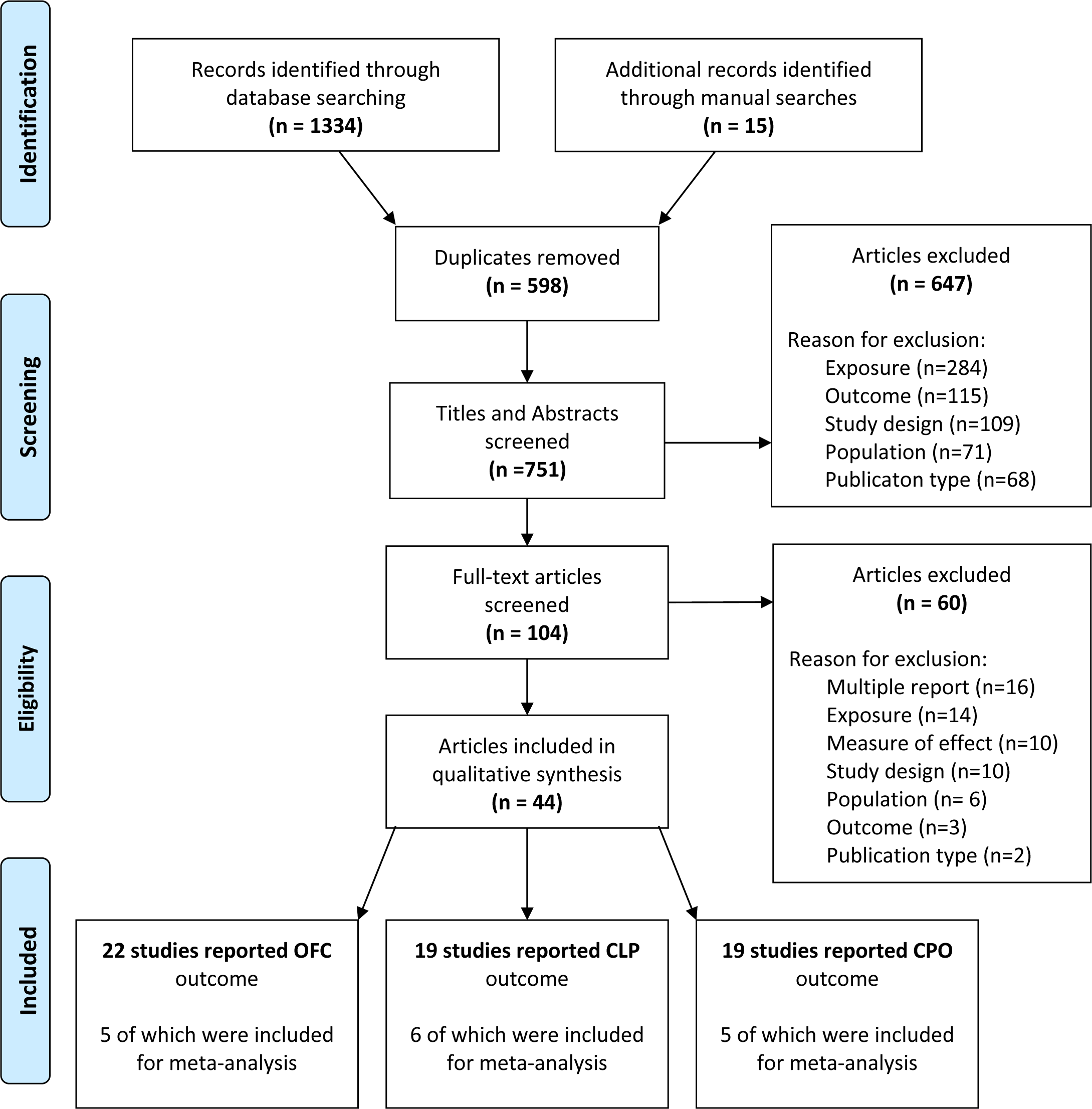
A flow chart of the search strategy and study selection

Titles and abstracts were reviewed independently by two reviewers (MF/KD) according to the specified inclusion/exclusion criteria and differences resolved through discussion to reach a consensus. Where an abstract was not available or where a decision on inclusion/exclusion could not be reached by reviewing the abstract alone, full text screening was similarly performed independently by two reviewers for inclusion and any disagreements resolved through discussion. When multiple reports of a study were identified, the study with the greatest number of patients was selected. The Rayyan web application was used to facilitate the screening process (Ouzzani et al., 2016).

### Data extraction

Data was extracted via Microsoft Forms into an excel spreadsheet. Data extracted included: title, authors, publication year, country of study population, study design, sample description, sample size, outcomes recorded, confounding factors measured and study outcomes including dose-response data. Adjusted measures of effect were extracted preferentially to reduce the impact of confounding factors. Data from each study was extracted by one reviewer (MF) and checked for accuracy by a second reviewer (KD)(Centre for Reviews and Dissemination, 2009).

### Assessment of study quality

The Newcastle-Ottawa Scale (NOS)(Wells et al., 2000) was used to assess the quality of cohort and case-control studies included in this systematic review. The NOS for cohort studies consists of eight questions amongst three domains (selection, comparability and outcome). Similarly, the NOS for case-control studies consists of eight questions amongst three domains (selection, comparability and exposure). Stars are awarded for adequate methodology and were used to allocate a score of good, fair or poor to each study with pre-defined criteria (see Supplementary Table 3). Good and fair studies were deemed appropriate for meta-analysis, whereas studies categorized as poor were deemed to be of too low quality for inclusion. Maternal age and maternal alcohol consumption were identified as the most important confounding factors, followed by folic acid supplementation and obesity, based on previous findings (Bille et al., 2005; Badovinac et al., 2007; Molina-Solana et al., 2013; Izedonmwen et al., 2015). Studies were required to adjust for maternal age and alcohol consumption in order to achieve at least a ‘fair’ rating and be included in the meta-analysis.

Funnel plots were used to visually assess the likelihood of small study publication bias if more than 10 studies were included and Egger’s test was calculated to quantify funnel plot asymmetry (Sterne et al., 2011).

### Data Synthesis

A descriptive summary and narrative analysis of the included studies was performed, alongside an indication of study quality, in accordance with published guidance (Popay et al., 2006). Heterogeneity of the included studies was analyzed by exploring the study characteristics and using the I^2^ statistic where sufficiently similar studies were meta-analyzed.

The quantitative impact of maternal smoking as a cause of orofacial clefting was investigated using meta-analysis techniques where studies met the quality criteria for inclusion and shared sufficient methodological homogeneity. The minimum number of studies to conduct a meta-analysis was two. Pooled estimates for binary outcomes were calculated using the inverse variance method. The odds ration (OR) was the principle summary measure extracted from the primary studies and meta-analyzed. The fixed effects model was used where levels of statistical heterogeneity were low (I^2^ <50%); otherwise the random effects model was used. The population attributable fraction (PAF) was calculated to assess the public health impact (Mansournia and Altman, 2018) using the pooled odds ratio and the prevalence of exposure among cases (Miettinen, 1974). The dose-response impact of maternal smoking was analyzed for studies in which the smoking dose categories used by the included studies were analogous. Subgroup meta-analysis of the smoking dose categories was performed using the random effects model. Meta-analysis was performed using the “meta” package (Harrer et al., 2021) via the R Project for Statistical Computing (http://www.R-project.org/).

## RESULTS

### Study Selection and Study Characteristics

A flowchart for the article review process is shown in Figure 1. A total of 1334 citation records were identified from searching the four databases. A manual search of relevant systematic reviews and included studies identified 15 additional studies. After exclusions (see Supplementary Table 4), 45 studies from 44 publications were included in the systematic review (one publication reported two case-control studies from distinctly separate populations (Shi et al., 2007); 11 cohort studies and 34 case control studies (see Table 1). In total, 28,405 mothers giving birth to a live born child with CL/P have had their smoking status during pregnancy analyzed amongst the 45 studies.

**Table 1:**
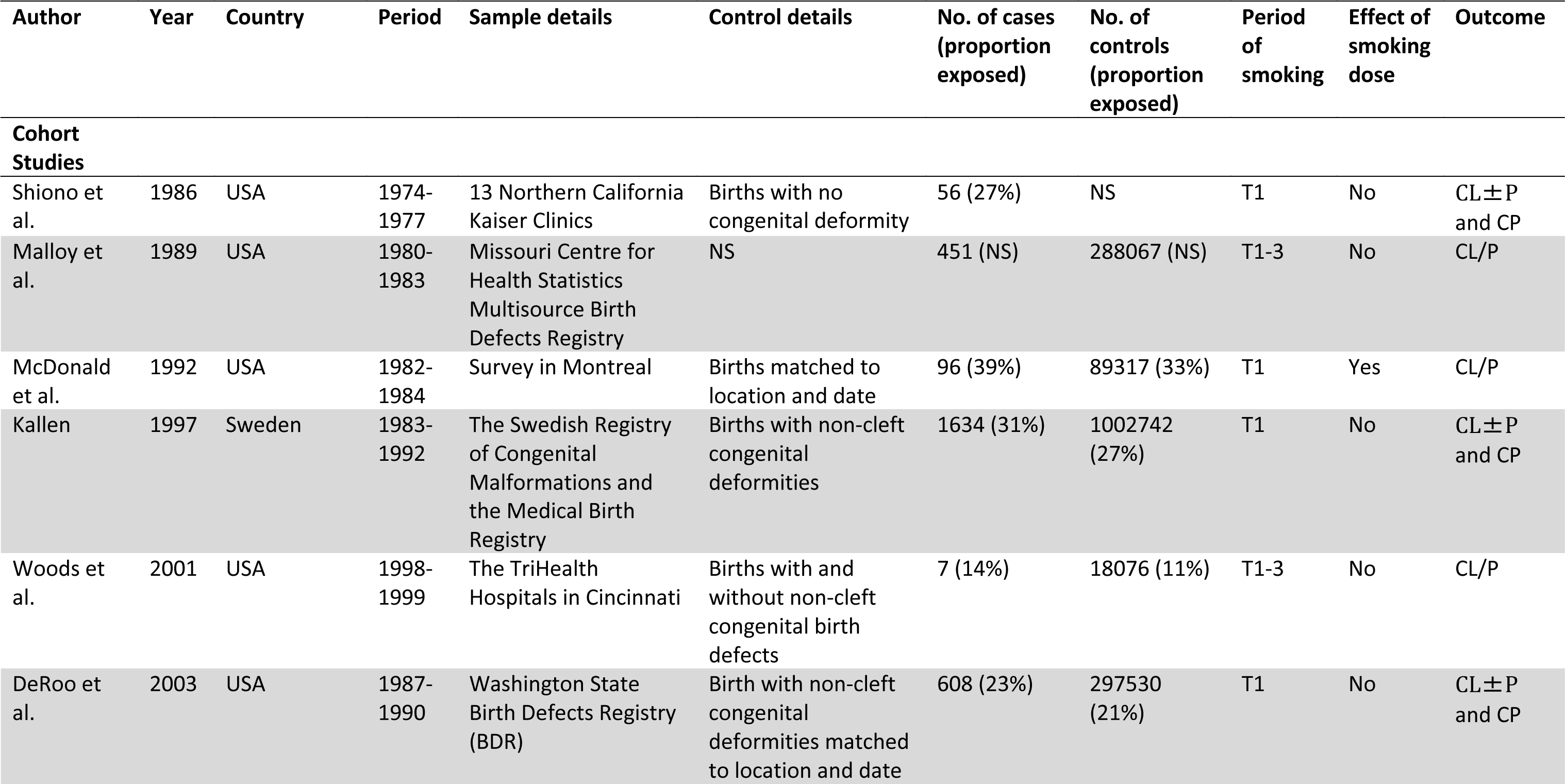
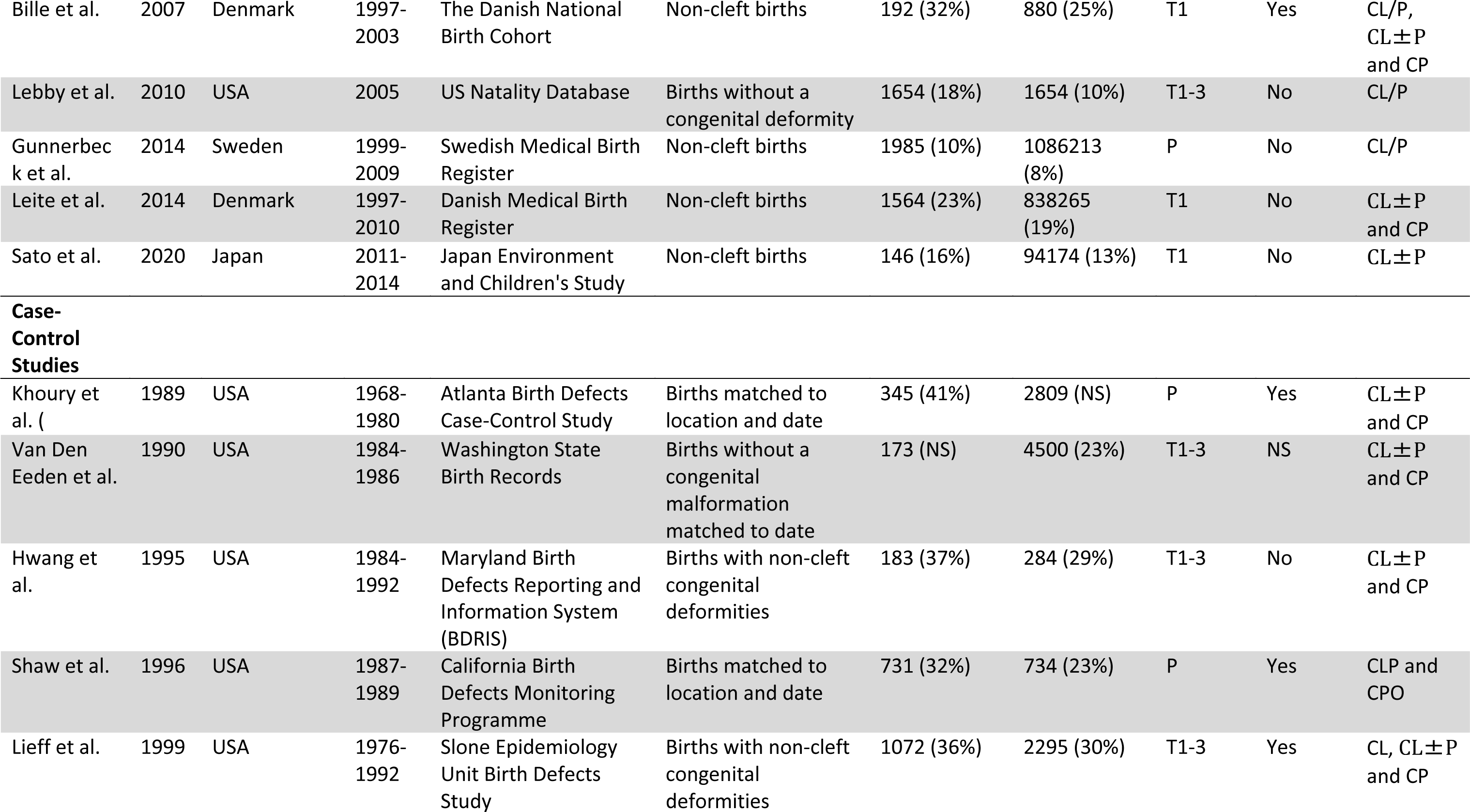
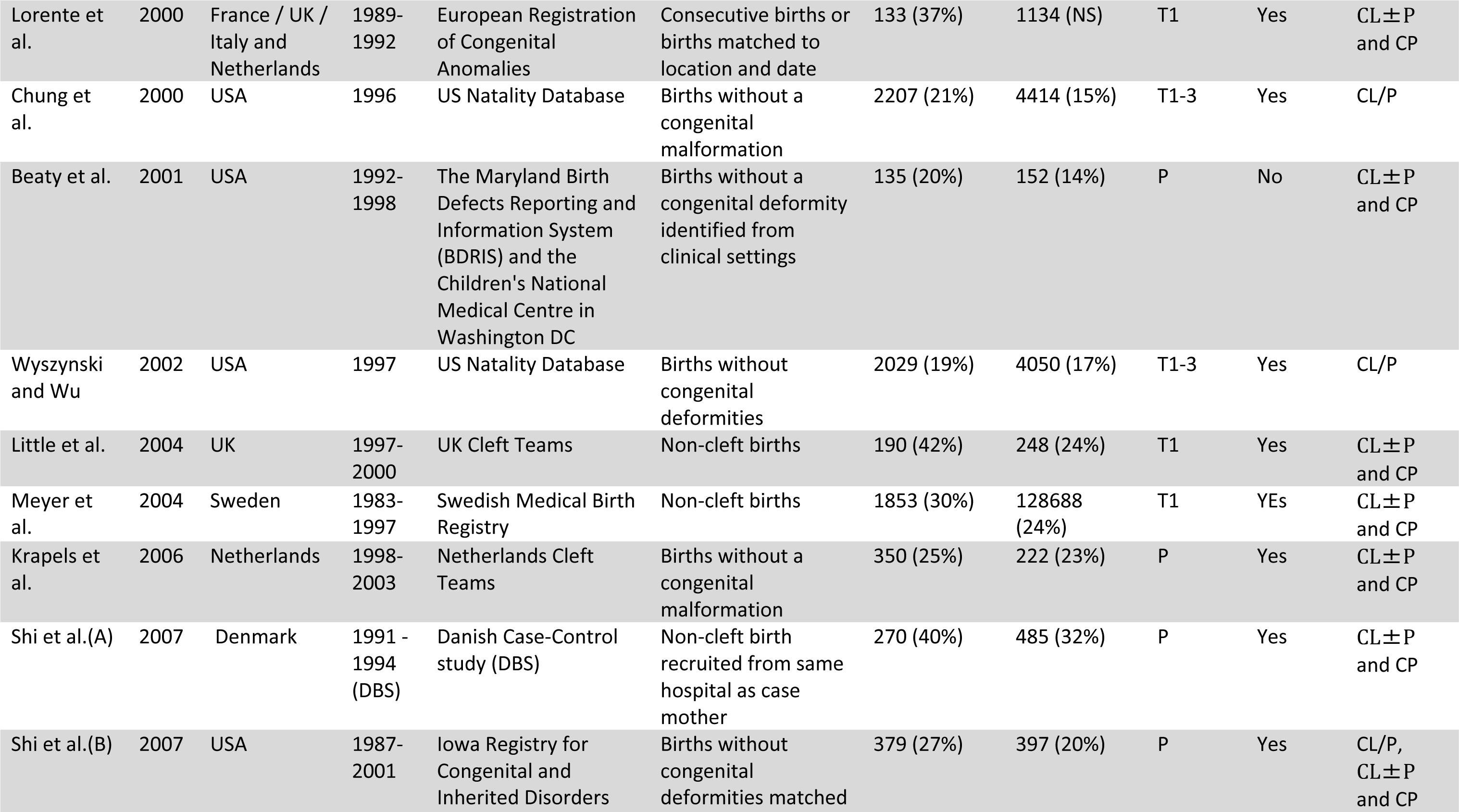
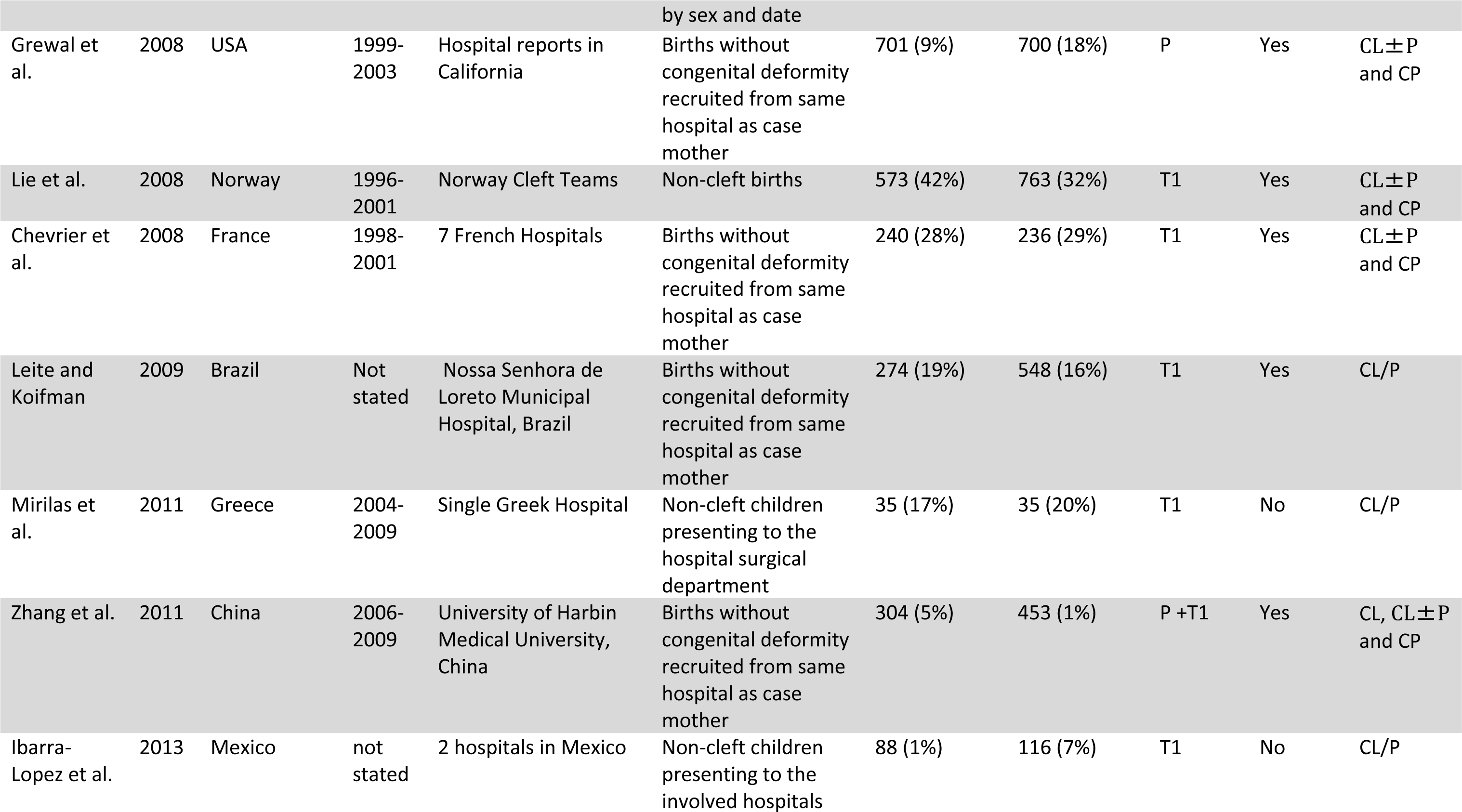
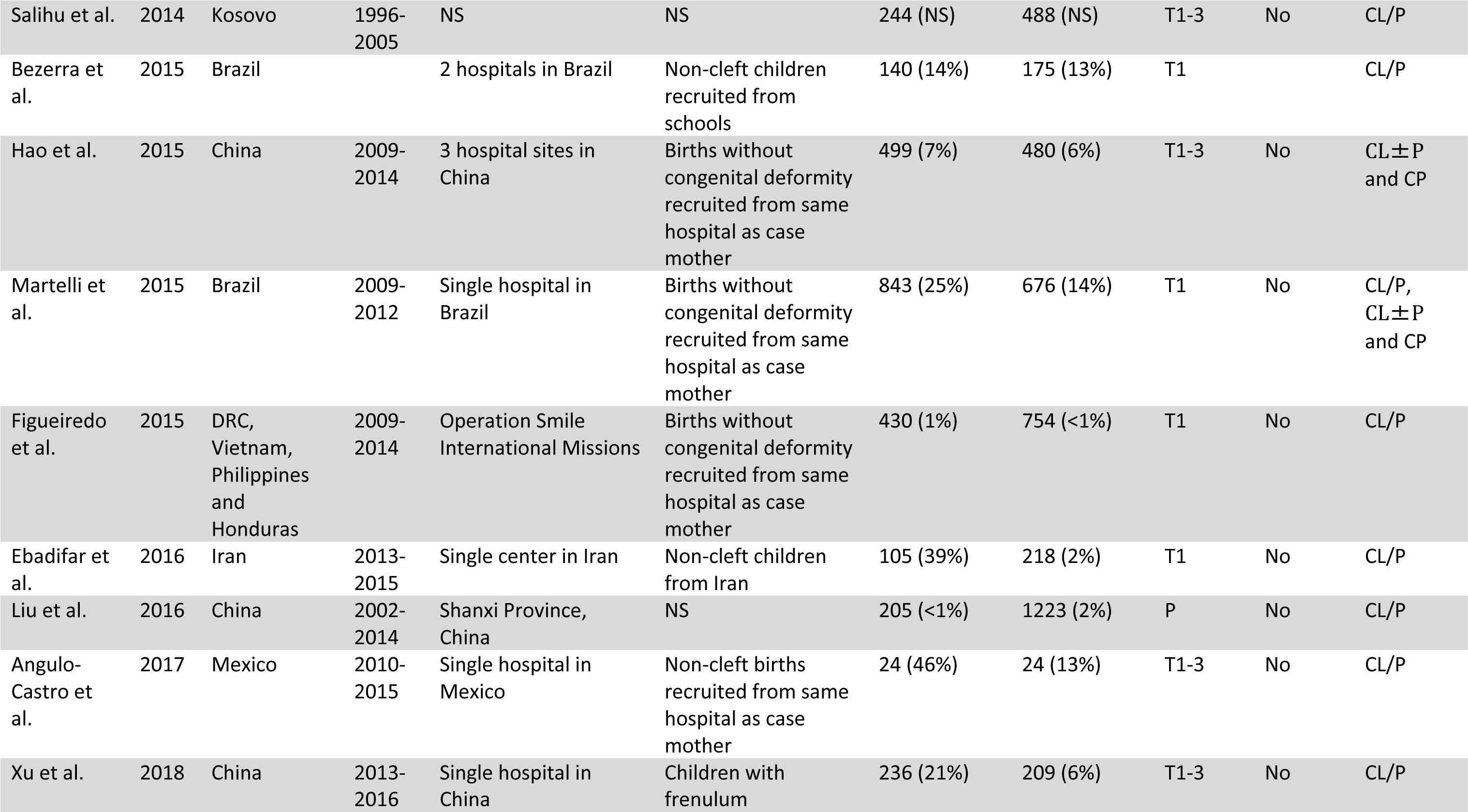
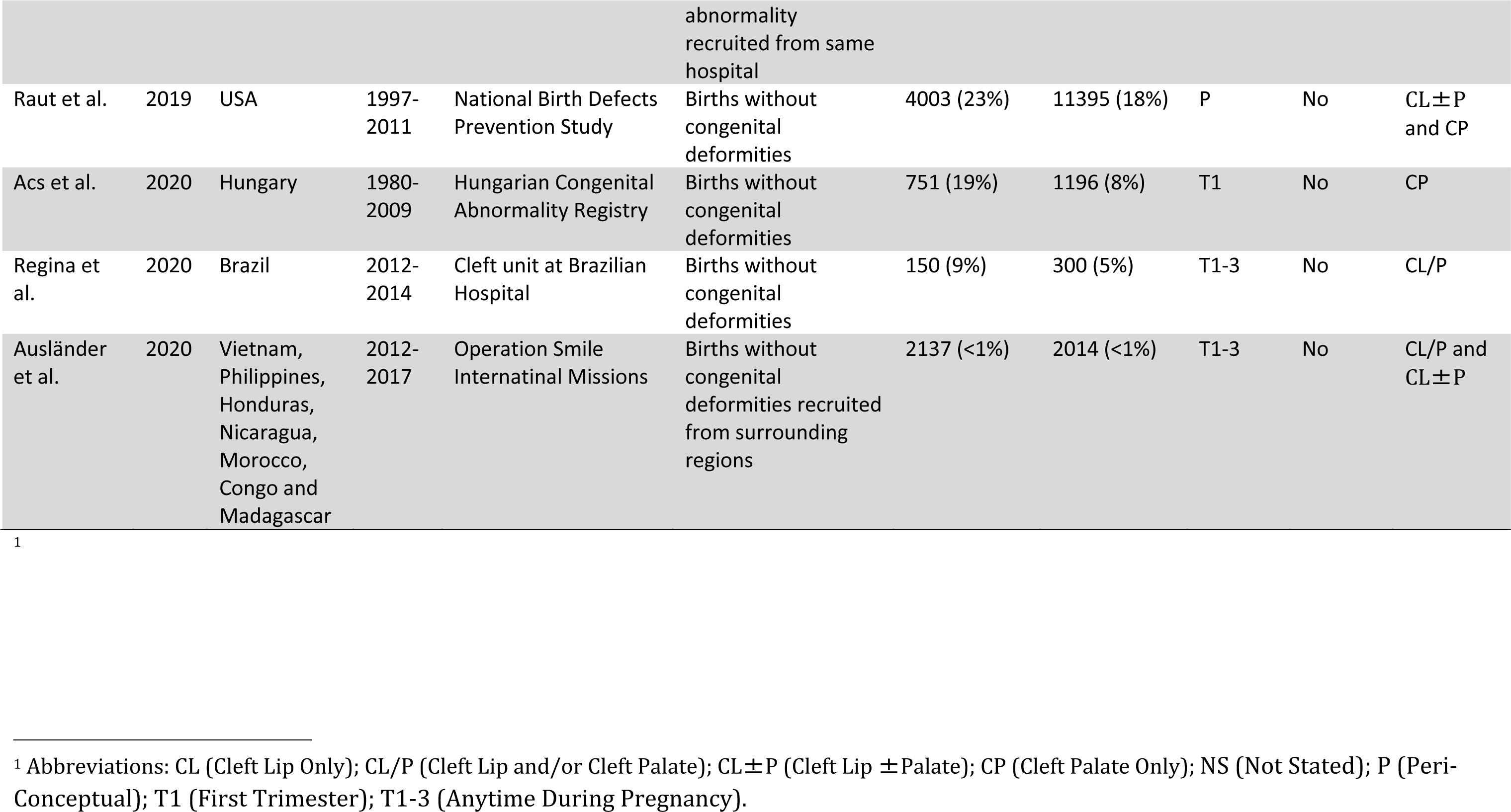
Characteristics of Included Studies.

### Reported outcomes

Twenty-two studies reported on CL/P outcome, with the funnel plot not indicating publication bias (see Supplementary Figure 5) and an Egger’s test of 0.77 (95% Confidence Interval (CI): 0.32, 1.85, *P*=0.18). Nineteen studies reported on CL±P outcome with the funnel plot not indicating publication bias and an Egger’s test of 0.09 (95% CI: -0.1, 1.17, *P*=0.88). Nineteen studies reported on CP outcome with the funnel plot not indicating publication bias and an Egger’s test of -0.28 (95% CI: -1.77, 1.2, *P*=0.71). As only two studies reported with cleft lip alone as the outcome, a funnel plot was not performed for these.

Nine studies reported smoking dose-response effects for CL/P outcome, a further 14 studies gave results by smoking dose for CL±P as the outcome and 13 studies for CP as the outcome.

Table 2 shows the study quality assessment for cohort and case-control studies based on the NOS. Only one study(Raut et al., 2019) of the 45 included studies had low scores in all eight NOS questions. Three studies were deemed to be good quality, 13 studies were deemed fair quality, and 29 deemed poor quality and the latter were excluded from the meta-analysis. A greater proportion of cohort studies (5/11) met the quality threshold for meta-analysis inclusion than case-control studies (11/34). The most common area lacking was the failure to adjust for confounding factors. The potential for exposure recall bias was present in all 34 of the case-control studies as by definition, information on exposure was collected retrospectively. Only four out of 11 cohort studies collected maternal smoking exposure data prospectively.

**Table 2:**
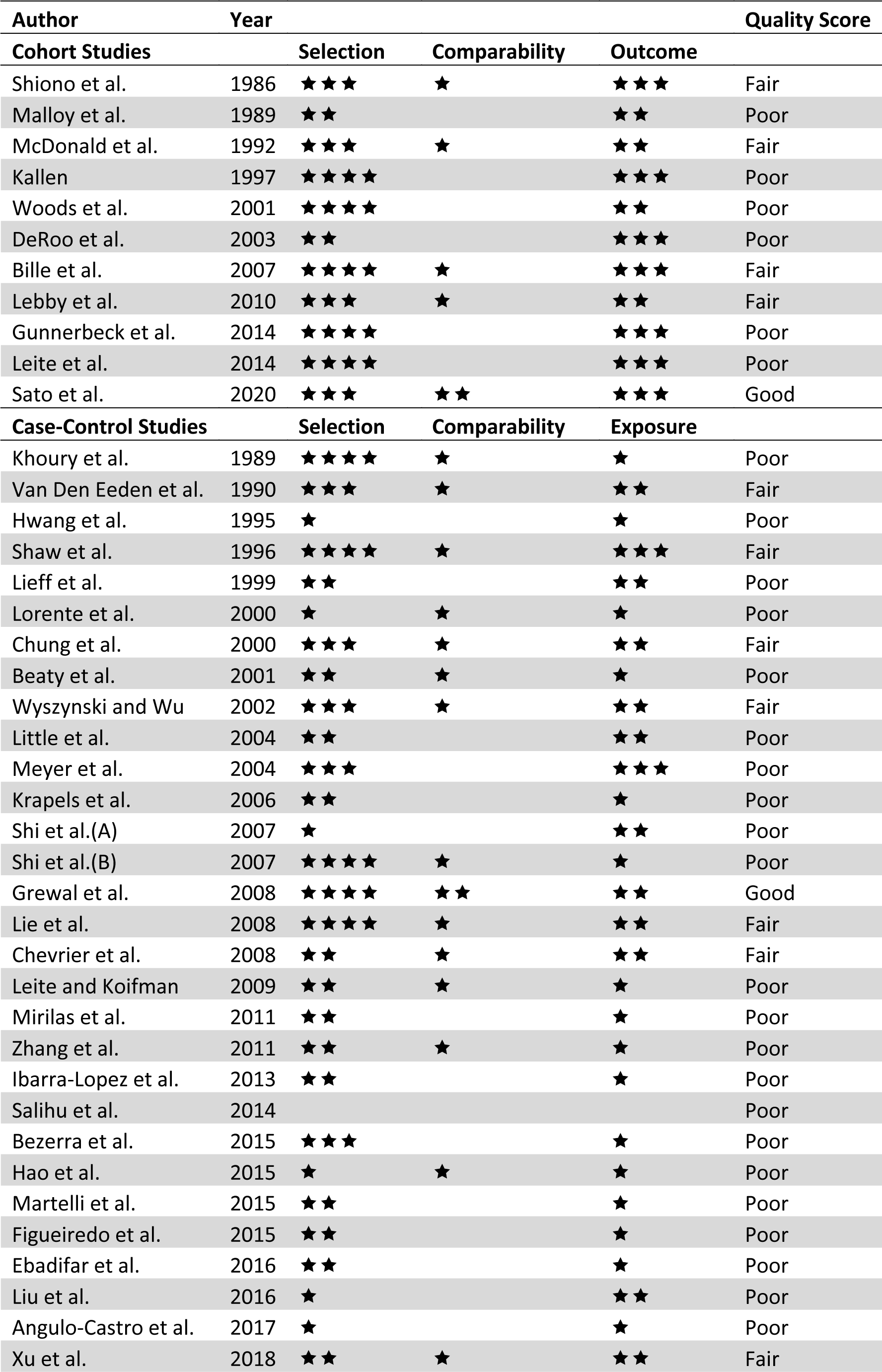
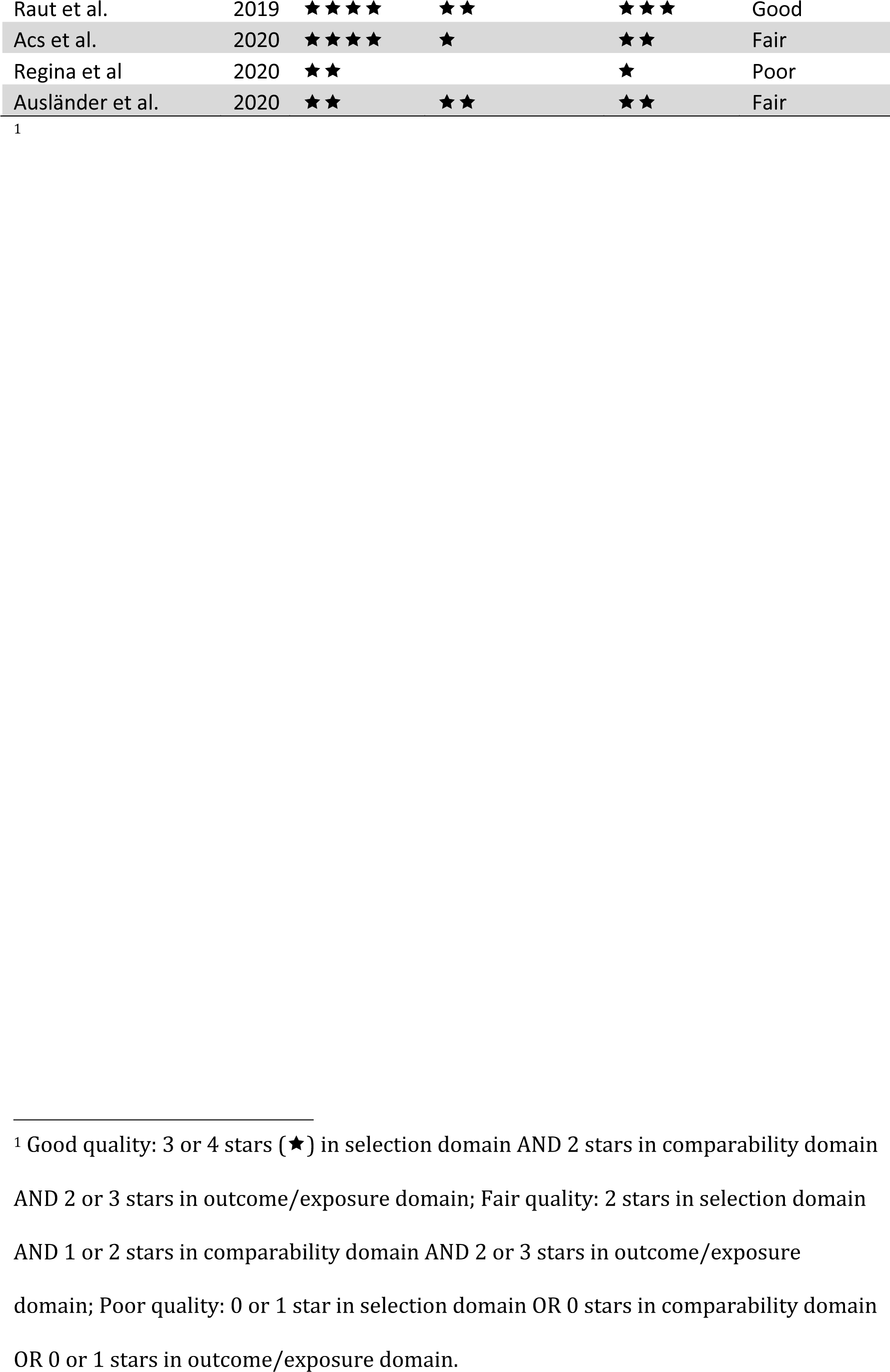
Quality Assessment of Included Studies using the Newcastle Ottawa Scale.

All of the 11 cohort studies were truly or somewhat representative of the general population and were able to demonstrate the outcome of interest was not present at the start of the study. Of the case control studies, 7 out of 34 did not meet the participant selection domain criteria due to failing to demonstrate independent validation of case definition (11 of 34), the potential for selection bias of cases (23 of 34) and/or selected controls from hospitalized populations (21/34).

Comparability criteria was not met in 6 out of 11 cohort studies and 20 out of 34 case-control studies due to not adjusting for at least maternal age and maternal alcohol consumption as confounders in the analysis. Folic acid supplementation and obesity were adjusted for in less than half of included studies (see Supplementary Table 5).

All of the 11 cohort studies used record linkage to verify OFC outcome. Exposure criteria was not met by 18 out of 34 case-control studies because of relying on self-assessment (8 of 34), using an interviewer who was not blinded to case/control status (23 of 34) and/or the non-response rate of cases/controls was not described (20 of 34).

### Meta-analysis

Five studies reporting effect estimates for smoking and CL/P were included in the meta-analysis (see Figure 2). There was no strong evidence of between study heterogeneity (I^2^=27%, *P*=0.24). The pooled OR using the fixed effects model was (95% CI: 1.27, 1.59). Based on the proportion of maternal smoking amongst case mothers of 14% in these five studies, the PAF was 4% (95% CI: 3%, 5%).

**Figure 2:**
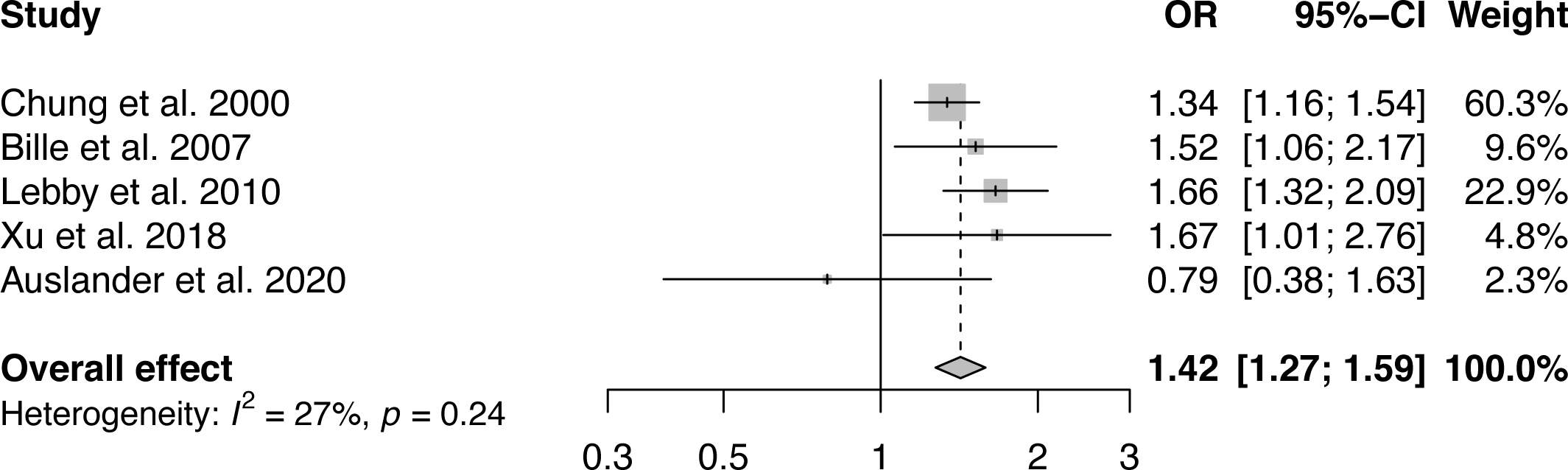
Forest plot to display the measures of effect for studies reporting cleft lip and/or palate outcome. The overall effect has been calculated using a fixed effects method.

Six studies reporting the effect for smoking and CL±P were included in the meta-analysis (see Figure 3). There was no evidence for statistical heterogeneity between the studies (I^2^ = 0%, *P*=0.67). The pooled OR using the fixed effects model was 1.31 (95% CI: 1.19, 1.45). Five studies reporting measures of effect for smoking and CP were included in the meta-analysis (see Figure 4). The statistical heterogeneity between the studies was high (I^2^ = 81%, *P*<0.01) due to an outlying case-control study performed in Hungary (Ács et al., 2020), reporting a stronger positive effect of smoking on CP than the other included studies. The pooled OR using the random effects model was 1.49 (95% CI: 1.01, 12.19). The exclusion of the outlying study in the CP meta-analysis resulted in no evidence for statistical heterogeneity (I^2^ = 0%, *P*=0.49) and a fixed effect pooled OR of 1.25 (95% CI: 1.09, 1.44). It was not possible to calculate the PAF for maternal smoking and CL±P or CP due to missing data in included studies, precluding calculation of prevalence of exposure.

**Figure 3:**
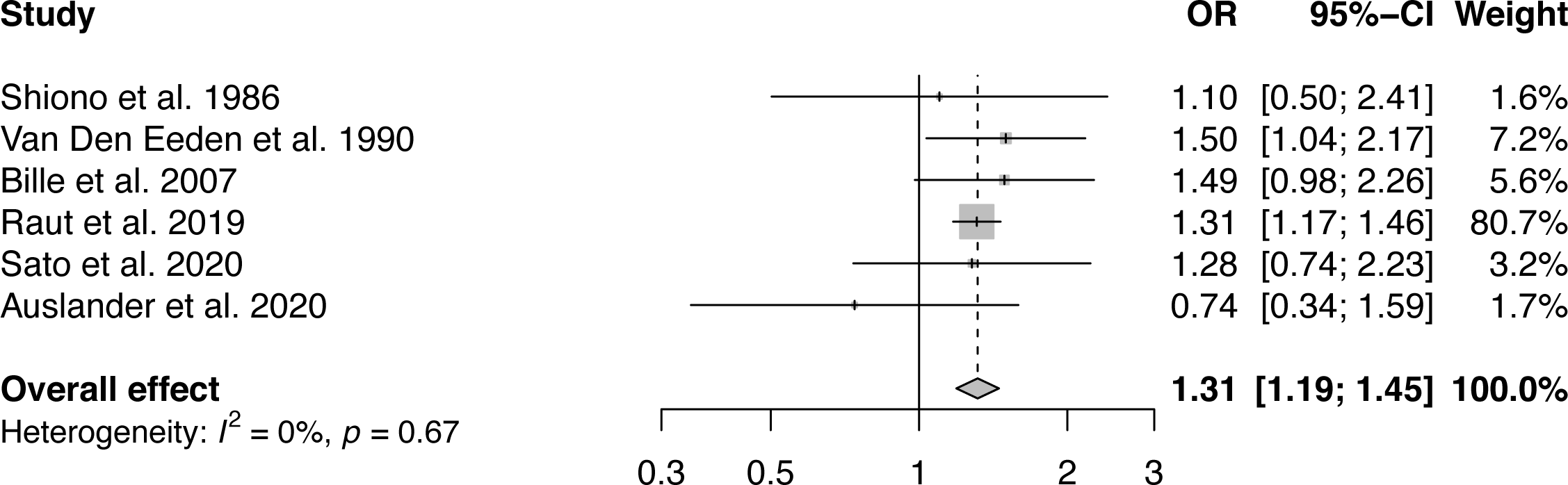
Forest plot to display the measures of effect for studies reporting cleft lip ± palate outcome. The overall effect has been calculated using a fixed effects method.

**Figure 4:**
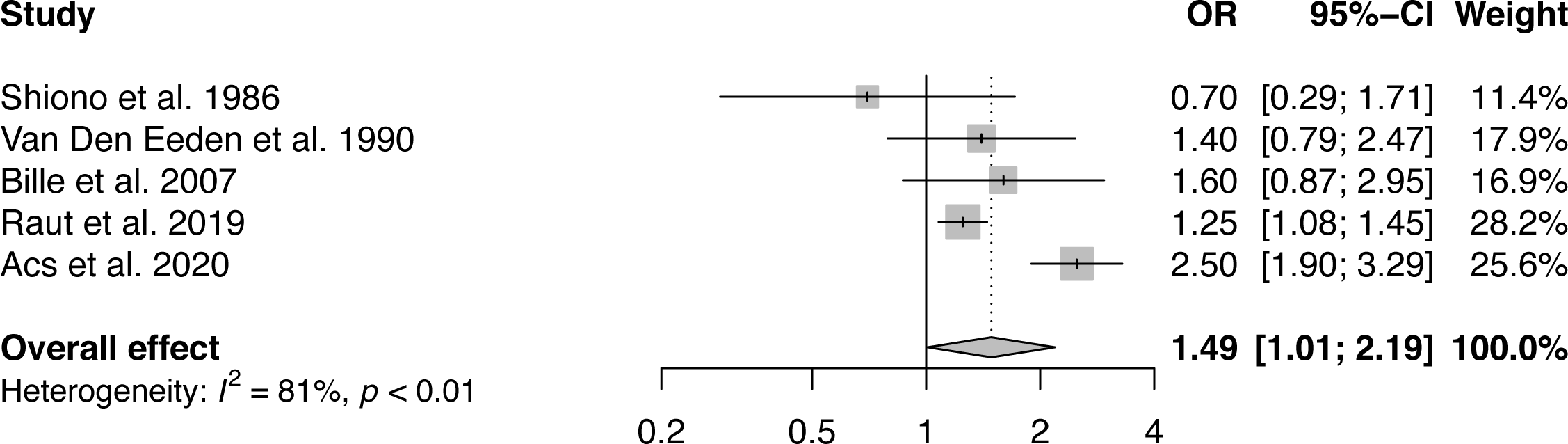
Forest plot to display the measures of effect for studies reporting cleft palate only outcome. The overall effect has been calculated using a random effects method.

Individual study effect estimates and pooled analysis for all studies included in this systematic review reporting outcomes for CL/P, CL±P and CP can be found in Supplementary Figures 6-8.

### Subgroup analysis

Five studies reporting measures of effect for the dose of smoking and CL/P were included in the subgroup meta-analysis (see Figure 5). All five studies measured three doses of smoking (low, medium and high) with comparable numbers of cigarettes smoked per day at each dose (1-10, 11-20 and >20 cigarettes per day). The pooled OR for the lowest dose of smoking was 1.20 (95% CI: 1.06, 1.36), for intermediate dose was 1.15 (95% CI: 0.97, 1.37) and highest dose was 1.45 (95% CI: 1.05, 2.00).

**Figure 5:**
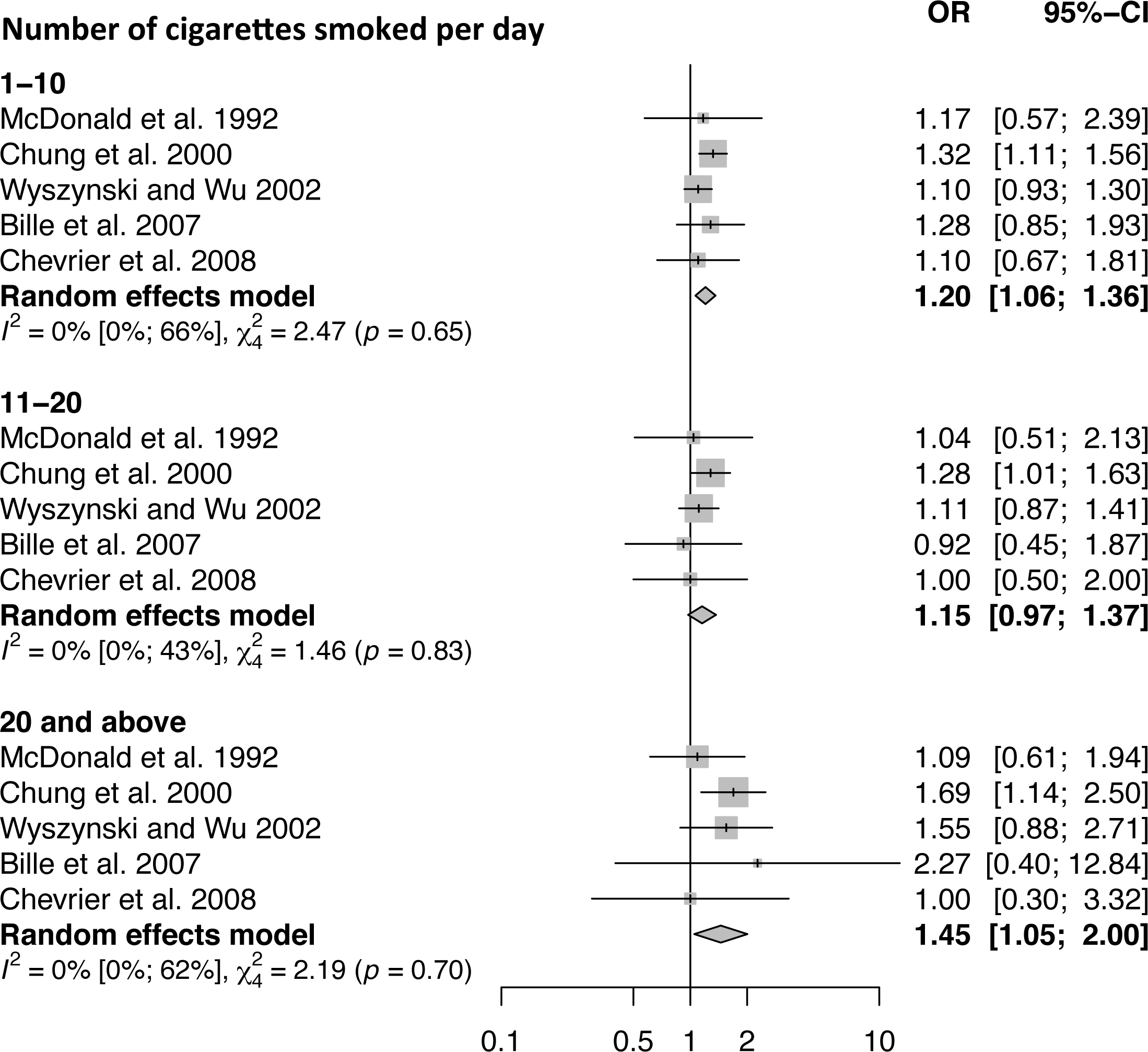
A subgroup forest plot to display the dose-response effect of smoking on cleft lip and/or palate outcome. The overall effect for each of the three dose categories (1-10, 11-20 and >20) has been calculated using a random effects model

Four studies were eligible for inclusion into the meta-analysis of the effect of smoking dose for both CL±P and CP respectively, but it was not possible to perform a meta-analysis because the reported smoking dose levels were not comparable.

## DISCUSSION

### Summary of evidence

There has been a large body of work to investigate the role of active maternal smoking in CL/P etiology, as shown by the 45 studies that met our inclusion criteria. This high volume of research should have provided a clear indication of the association between maternal smoking and CL/P, but the poor quality of studies overall has compromised the reliability of the reported findings. Only three studies out of the 45 included in this review were judged to be of good quality (Grewal et al., 2008; Raut et al., 2019; Sato et al., 2020;). The most common reason for poor quality within the studies was a failure to adjust for recognized confounding factors, placing the analyses at high risk of bias. Mother’s age, alcohol intake and obesity are all strongly associated with smoking behavior and all have been hypothesized to be risk factors for orofacial clefts. Furthermore, alcohol intake during pregnancy is a known teratogen, making the adjustment of these confounding risk factors even more critical (Carreras-Torres et al., 2018; Taylor et al., 2018; Taylor et al., 2019).

Our meta-analysis suggests that maternal smoking may have a moderate role in CL/P etiology with pooled OR of 1.42 (95% CI: 1.27, 1.59). The PAF estimates the proportion of the disease that would be reduced by eliminating exposure to a given risk factor, assuming the risk factor is causal. Smoking has previously been found to account for the largest risk when PAFs are calculated for a number of modifiable risk factors for CL/P (Raut et al., 2019). The pooled PAF of 4% (95% CI: 3%, 5%) in this review is similar to the previously reported range of 4-6% from three individual studies (Honein et al., 2007; Honein et al., 2014; Raut et al., 2019;). The indication here is that should maternal smoking be eliminated, 4% of CL/P would not occur. Evidence of a dose-response relationship can add support to a causal relationship. The analysis of dose effect in CL/P demonstrated the highest dose of smoking (>20 cigarettes per day) to have the strongest positive effect on risk of cleft, but the intermediate smoking dose (11-20 cigarettes per day) had a similar effect to the lowest dose (1-10 cigarettes per day). This may represent a threshold effect of more than 20 cigarettes needing to be smoked a day before a difference is noted in CL/P etiology. Alternatively, the greater effect in the highest smoking dose may reflect the propensity for risk taking behaviors associated with additional confounding by substance abuse (such as alcohol), which may not have been adequately adjusted for. The effect of the highest smoking dose on CL/P etiology should be interpreted with caution as the number of cases within the individual studies were less than for low and medium smoking doses, therefore the effect estimates were less precise.

Historically, CL/P has been subdivided in to CL±P and CP, reflecting different embryological origins from the primary palate and secondary palate respectively (Dixon et al., 2011). Studies included in this review reported individual outcomes for CL±P and CP and the respective pooled ORs demonstrated a moderately positive association with maternal smoking, similar to that of OFC. The pooled OR for CP (OR = 1.49) was greater than for CL±P (OR = 1.31) and this is an inverse of the relationship reported in two previous meta-analyses (Little et al., 2004; Xuan et al., 2016). The pooled OR for CP reported in this review should be interpreted with caution as it was influenced by the outlying result of a single study (Ács et al., 2020), with a heterogeneity between studies present. The only study with a good quality rating included in the CP meta-analysis (Raut et al., 2019), reported a more modest measure of effect, therefore the pooled OR following exclusion of the outlying study (OR = 1.25) may be a more accurate representation of the effect of smoking on CP etiology.

### Strengths and Limitations

Strengths of this review include a comprehensive search strategy with concerted efforts made to include all languages and a wide variety of study designs. Thorough assessment of study quality facilitated the inclusion of studies into the meta-analysis only if they met pre-defined threshold criteria.

The main limitation of interpreting the results from the meta-analysis relate to the inherent flaws of the standard analytical cohort and case-control approaches and their associated potential for bias. Studies were included in the meta-analysis if they had adjusted for a minimum set of confounders (maternal age and maternal alcohol consumption), which means that there was scope for additional important confounding factors to be unaccounted for. Even when adjustment for all relevant confounding factors is performed, bias may be present due to inaccurate measurement of confounding factors, misclassifications of exposure and differential missing data (Lawlor et al., 2016). The small sample sizes of some studies included in the meta-analysis meant their effect estimates were imprecise. A dose-response relationship could not be tested in CL±P and CP outcomes due to differences in smoking dose categorization reported in the included studies. Restriction of the search to published studies could have introduced publication bias, despite the evidence for publication bias being weak. This review focused upon active cigarette smoking in females and whilst the association of both passive and paternal smoking on CL/P has been reported, there has been less scientific focus in these areas when compared to active maternal smoking (Savitz et al., 1991; Krapels et al. 2008; Figueiredo et al. 2015; Hao et al. 2015; Sabbagh et al. 2015).

### Interpretation

Our understanding of the causal role of maternal smoking in CL/P is limited because of biases affecting traditional observational methods and the impracticalities of performing randomized controlled trials in this setting. If our reported moderate association is an accurate reflection of the role that maternal smoking plays then we would predict that the elimination of this risk factor would result in the reduction of 8,000 less cases per year worldwide as it is estimated that 200,000 children are born with CL/P per year (Mossey et al., 2009; The Central Intelligence Agency, 2021). This estimation is based on a 14% prevalence of maternal smoking in case mothers, originating from high income country publications, whereas the World Health Organisation estimates 17% of the global population use tobacco products, mostly from low and middle income countries (World Health Organisation, 2020).

The potential for maternal smoking to play a moderate role in CL/P etiology fits within our current understanding about the cause of CL/P being complex, multifactorial and involving both environmental and genetic factors (Dixon et al. 2011). Gene-environment interactions between smoking and CL/P have been the focus of a number of studies over the last two decades and these have improved our understanding of the pathogenesis of CL/P (Vieira, 2008; Krapels et al., 2008; Beaty et al., 2016; Garland et al., 2020). If smoking only accounts for 4% of the population attributable fraction, the environmental and genetic factors accounting for the remaining 96%, and the interplay between them, remains to be defined.

### Recommendations / implications for practice/policy/ further research

This review seeks to address an important public health question regarding the role of maternal smoking in CL/P etiology. Tobacco use is still common worldwide in pregnancy and is the focus of campaigns by the World Health Organisation to reduce adverse health effects on woman and infants (World Health Organisation, 2013). The neonatal health risk associated with maternal smoking were highlighted to the public in 2014 by the U.S Surgeon General’s Report, with smoking reported to increase the risk of CL/P by 30-50% (United States Department of Health and Human Services, 2014). Focus group research has highlighted the difficulties of changing smoking behaviors in pregnant women but suggests educational information with pictorial representation of babies risk may be an effective motivational method (Levis et al., 2014).

The methodologies used by the 45 eligible studies were all conventional observational design (cohort or case control designs). To strengthen our understanding of the causal role of maternal smoking in CL/P, this review highlights the need for high quality studies using a variety of methodological approaches with different directions of bias (Pearce et al., 2019). An instrumental variable model using genetic variants as proxies for smoking has been used in the past to assess the effect of maternal smoking on CL/P risk and reported a substantially stronger positive effect than traditional analytic studies, but the genetic variants used were not strongly associated with smoking and the sample size was small (Wehby et al., 2011). More powerful studies, using multiple novel epidemiological designs that can overcome some of the limitations of traditional methods are required and have been used as part of a triangulated approach to further the understanding of the causal role of cigarette smoking for other health outcomes (Gage et al., 2020).

## Data Availability

Secondary data published in the public domain used for this systematic review

## Acknowledgements

The authors thank Emma Place, Bristol Dental School Librarian, for her help with this study

## Abbreviations

CI: Confidence Interval
CL±P: Cleft Lip ± Palate
CP: Cleft Palate Only
NOS: Newcastle Ottawa Scale
CL/P: Cleft lip and/or cleft palate
OR: Odds Ratio
PAF: Proportional Attributable Fraction

## Supplementary Figure Legends

**Supplementary Figure 1:**
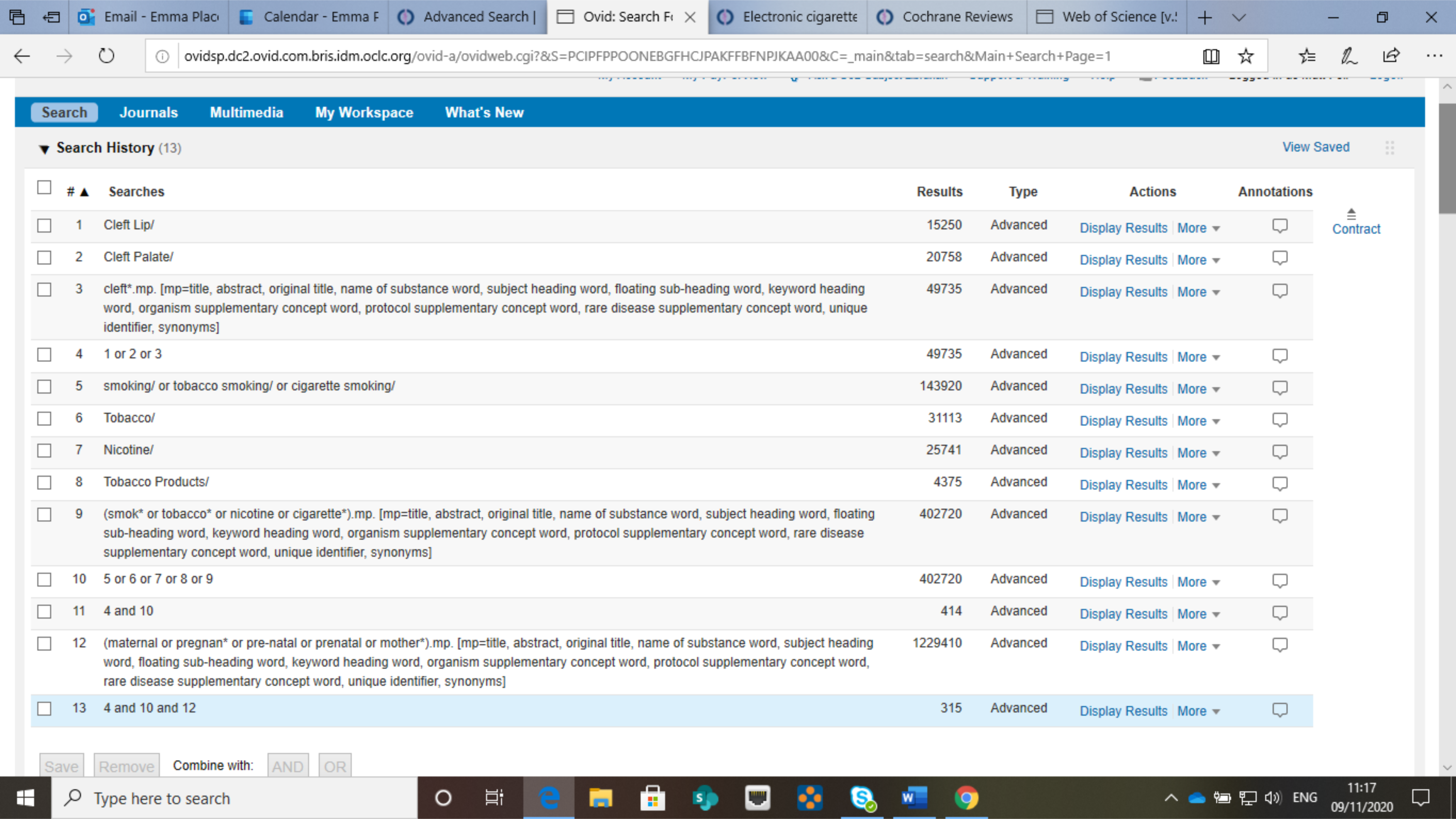
Medline search strategy

**Supplementary Figure 2:**
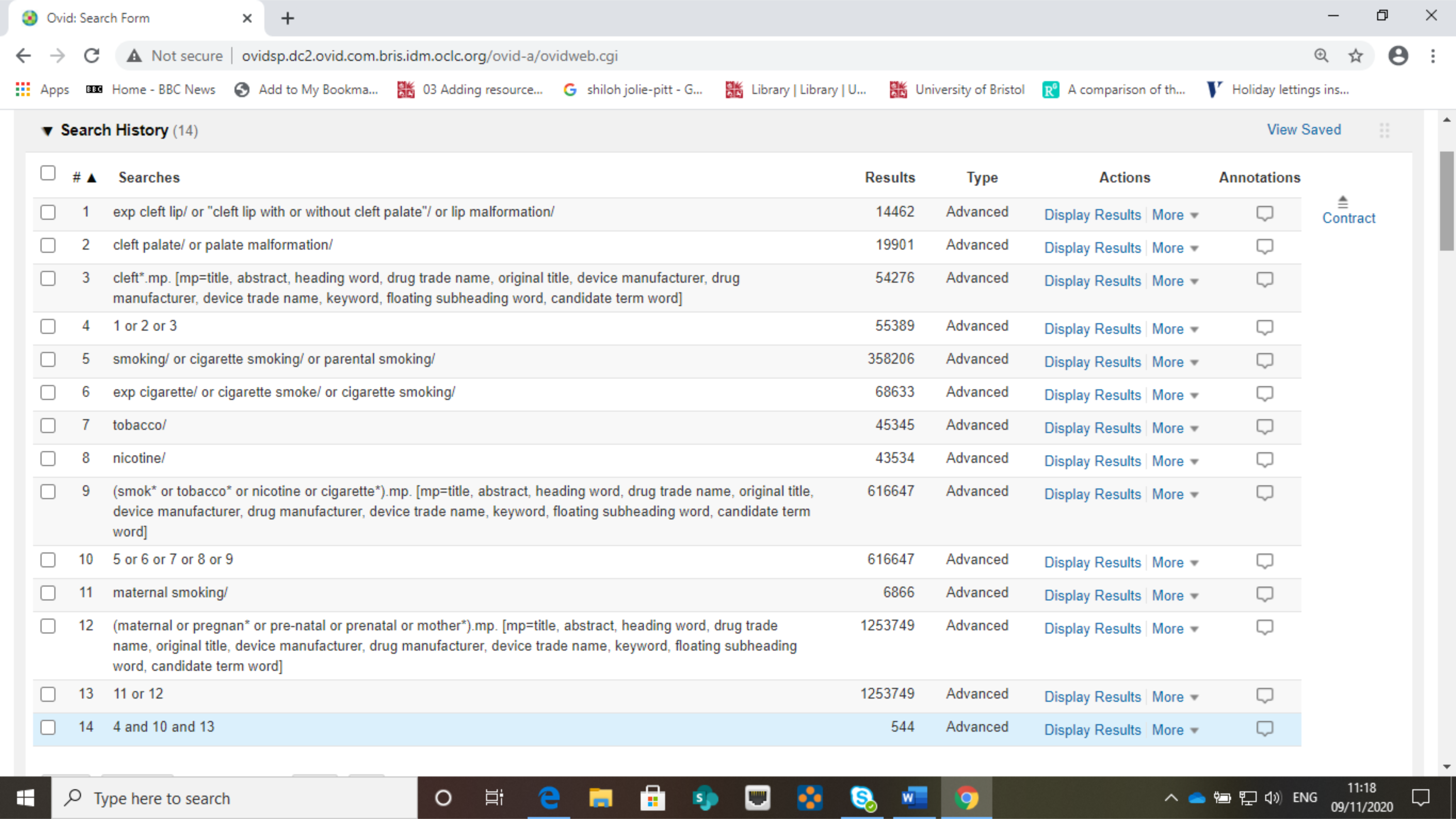
Embase search strategy

**Supplementary Figure 3:**
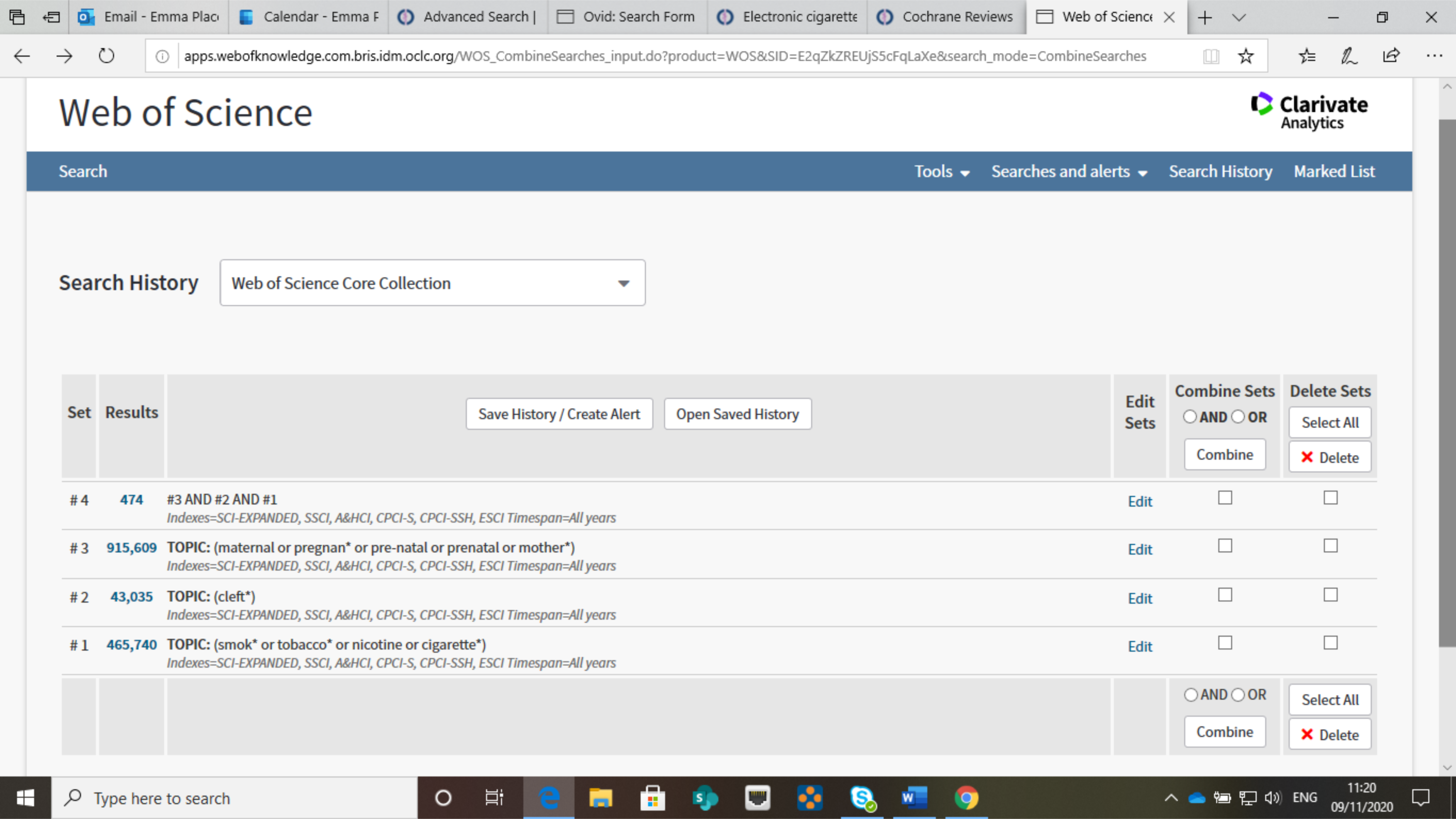
Web of Science search strategy

**Supplementary Figure 4:**
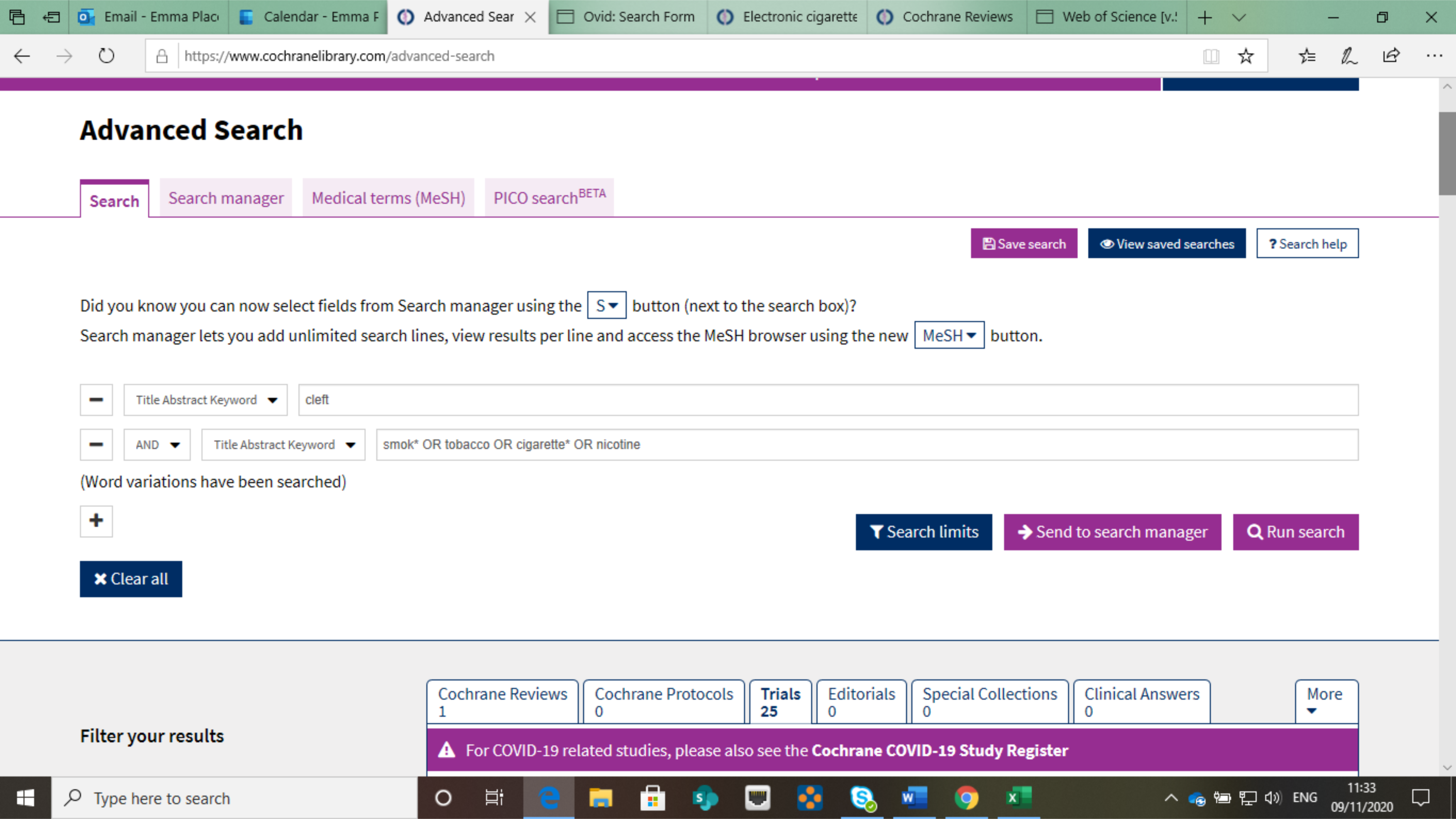
Cochrane search strategy

**Supplementary Figure 5:**
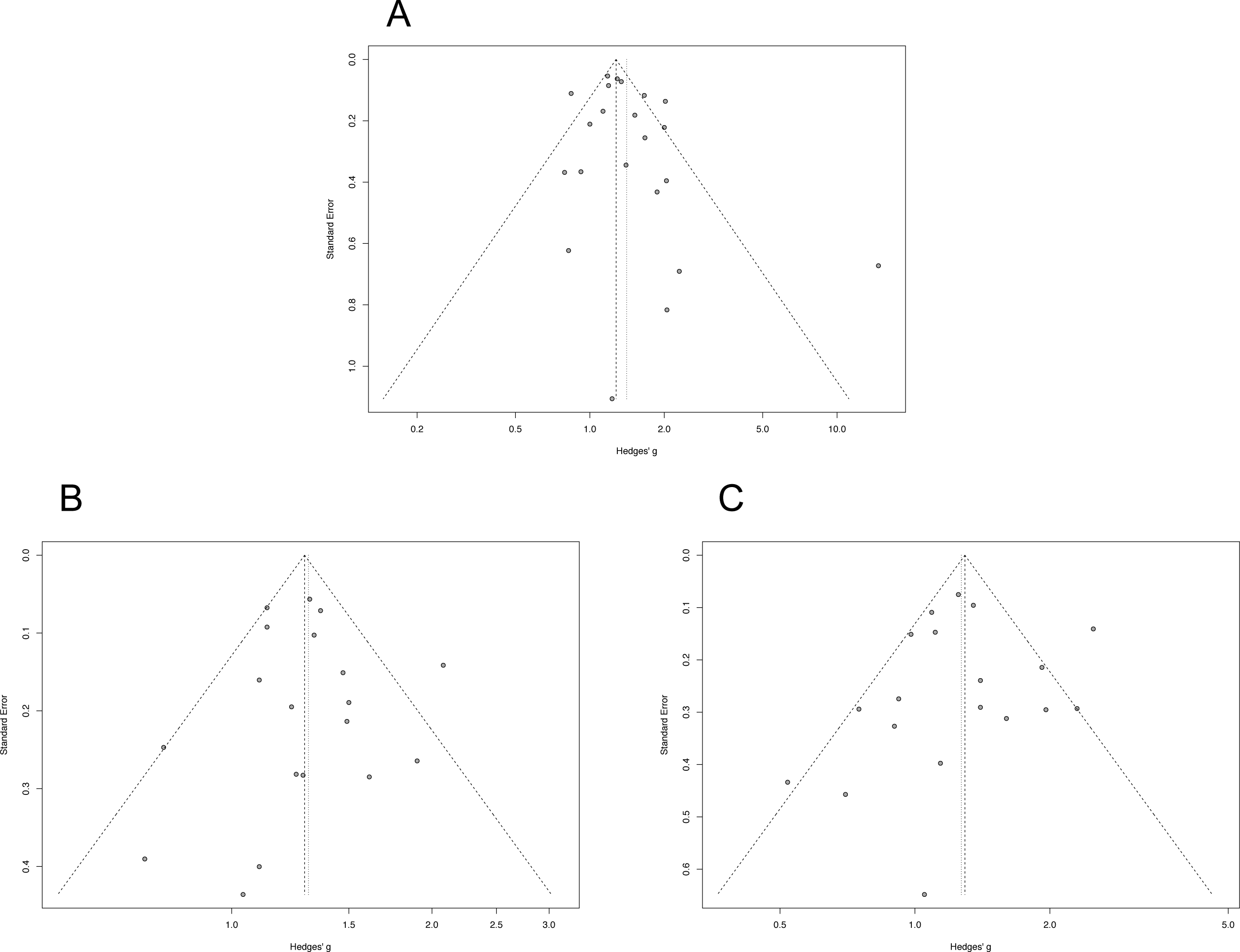
Funnel plots to test publication bias for studies included in this review. The studies have been categorized depending the outcome reported: A) Cleft lip and/or palate; B) Cleft lip ± palate; C) Cleft Palate Only

**Supplementary Figure 6:**
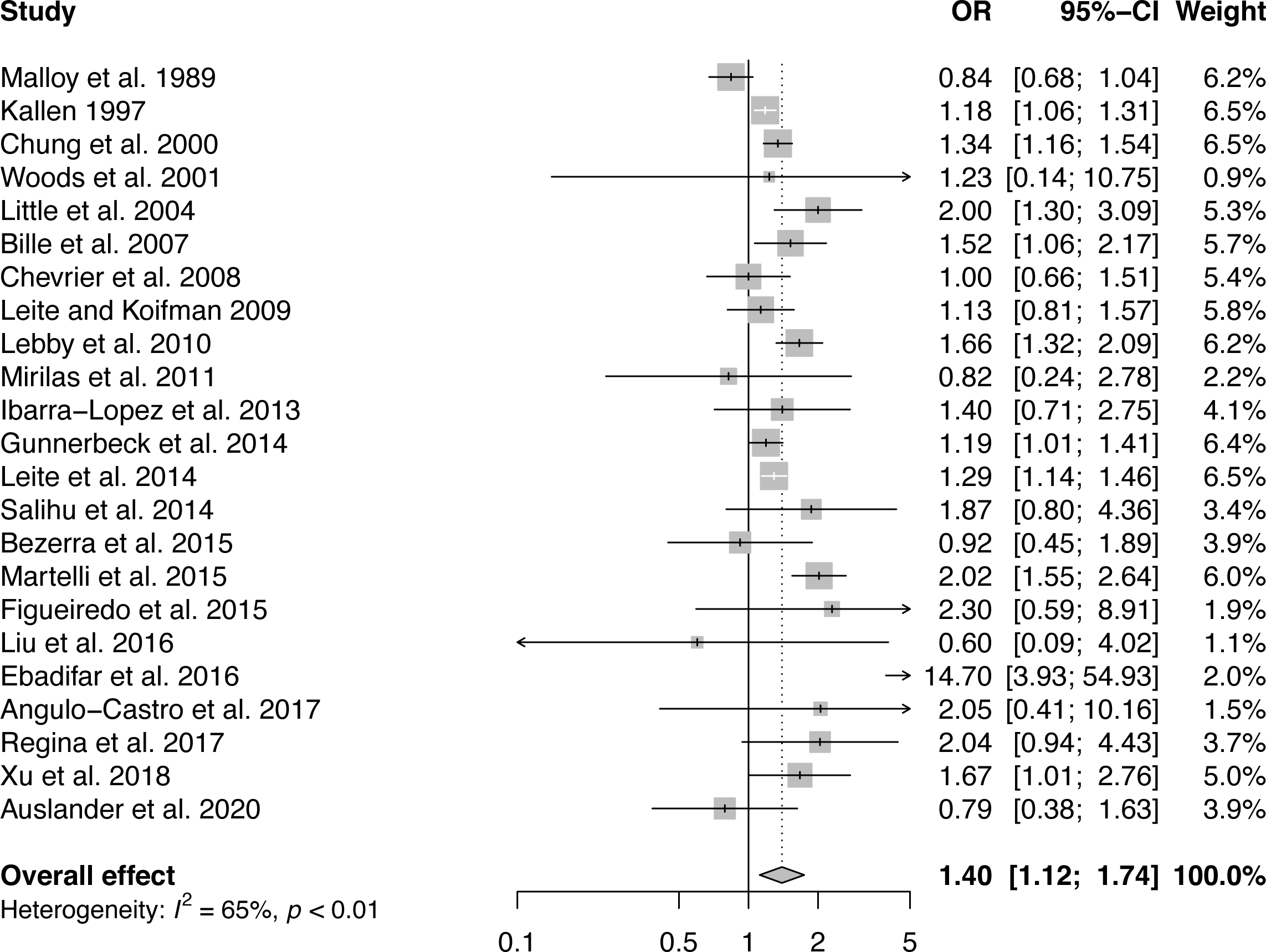
Forest plot to display the individual study measures of effect and pooled analysis for all studies included in this review reporting cleft lip and/or palate outcome.

**Supplementary Figure 7:**
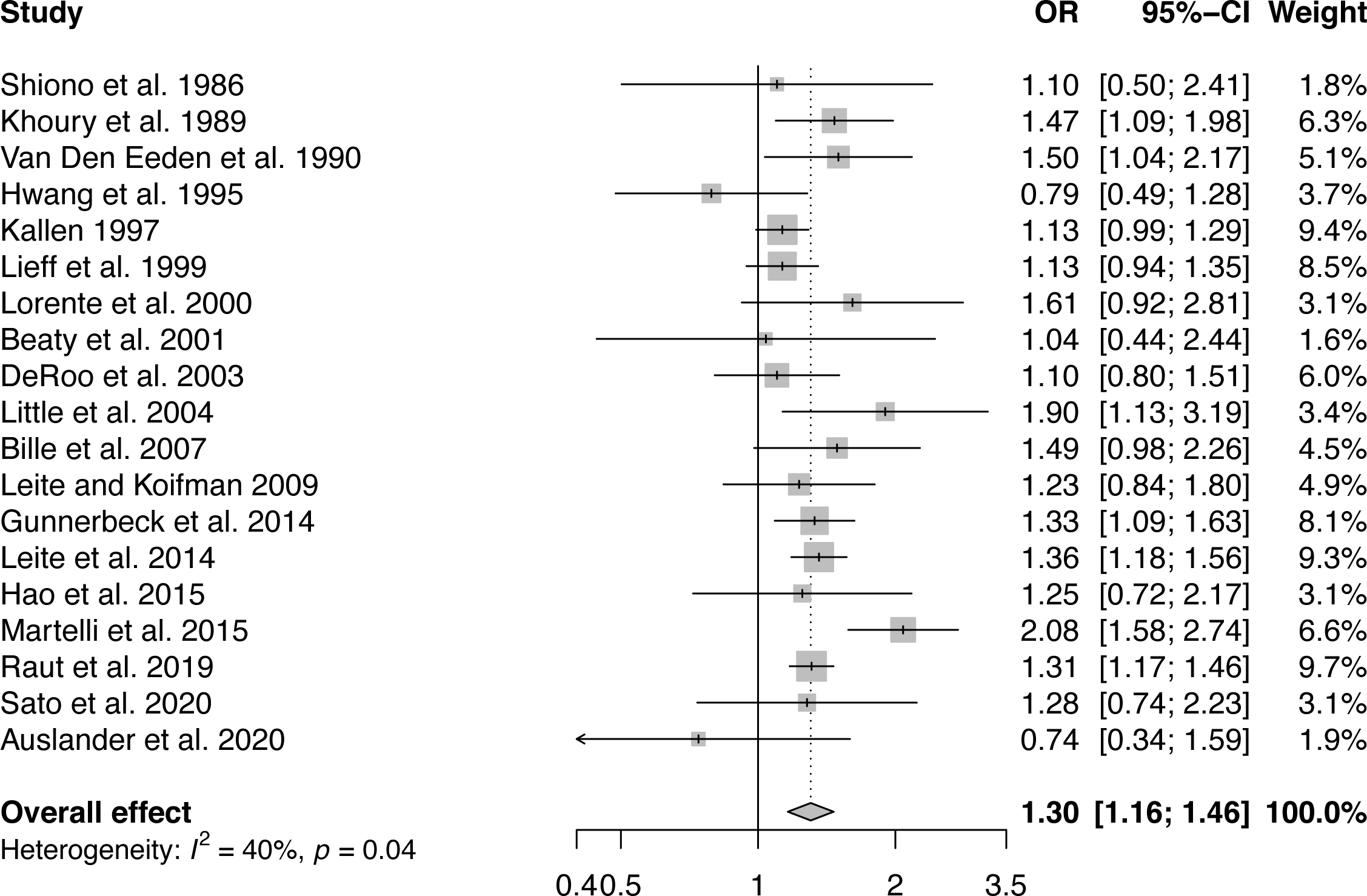
Forest plot to display the individual study measures of effect and pooled analysis for all studies included in this review reporting cleft lip ± palate outcome.

**Supplementary Figure 8:**
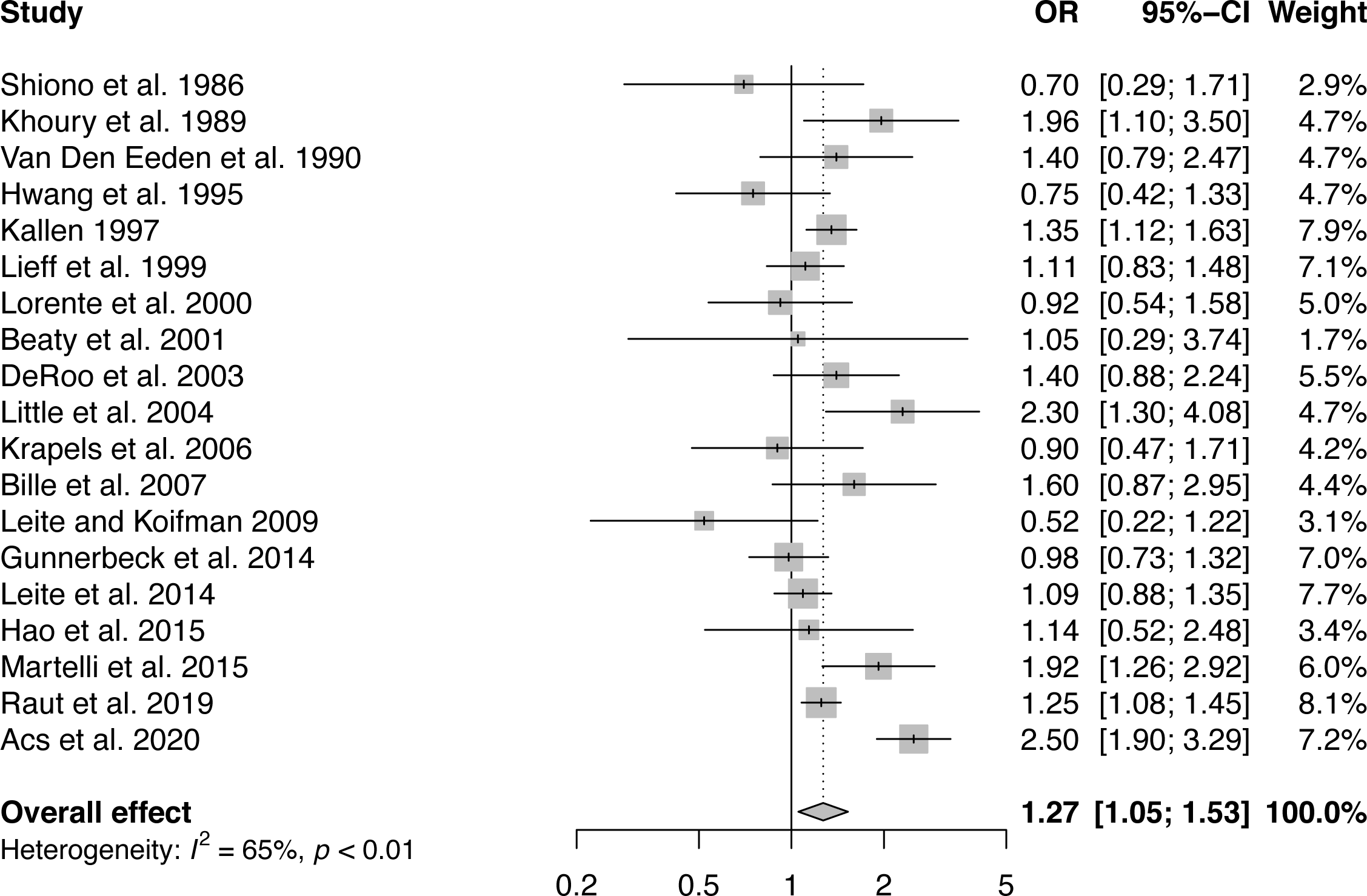
Forest plot to display the individual study measures of effect and pooled analysis for all studies included in this review reporting cleft palate only outcome.

**Supplementary Table 1:** PRISMA 2009 Checklist for the systematic review and metaanalysis

| Section/topic | # | Checklist item | Reported on page # |
| --- | --- | --- | --- |
| **TITLE** |  |  |  |
| Title | 1 | Identify the report as a systematic review, meta-analysis, or both. | 1 |
| **ABSTRACT** |  |  |  |
| Structured summary | 2 | Provide a structured summary including, as applicable: background; objectives; data sources; study eligibility criteria, participants, and interventions; study appraisal and synthesis methods; results; limitations; conclusions and implications of key findings; systematic review registration number. | 3 |
| **INTRODUCTION** |  |  |  |
| Rationale | 3 | Describe the rationale for the review in the context of what is already known. | 5 |
| Objectives | 4 | Provide an explicit statement of questions being addressed with reference to participants, interventions, comparisons, outcomes, and study design (PICOS). | Suppl. Table 2 |
| **METHODS** |  |  |  |
| Protocol and registration | 5 | Indicate if a review protocol exists, if and where it can be accessed (e.g., Web address), and, if available, provide registration information including registration number. | 5 |
| Eligibility criteria | 6 | Specify study characteristics (e.g., PICOS, length of follow-up) and report characteristics (e.g., years considered, language, publication status) used as criteria for eligibility, giving rationale. | 6 |
| Information sources | 7 | Describe all information sources (e.g., databases with dates of coverage, contact with study authors to identify additional studies) in the search and date last searched. | 7 |
| Search | 8 | Present full electronic search strategy for at least one database, including any limits used, such that it could be repeated. | Suppl. Figures 1-4 |
| Study selection | 9 | State the process for selecting studies (i.e., screening, eligibility, included in systematic review, and, if applicable, included in the meta-analysis). | 7 |
| Data collection process | 10 | Describe method of data extraction from reports (e.g., piloted forms, independently, in duplicate) and any processes for obtaining and confirming data from investigators. | 8 |
| Data items | 11 | List and define all variables for which data were sought (e.g., PICOS, funding sources) and any assumptions and simplifications made. | 8 |

|  |  |  |  |
| --- | --- | --- | --- |
| Risk of bias in individual studies | 12 | Describe methods used for assessing risk of bias of individual studies (including specification of whether this was done at the study or outcome level), and how this information is to be used in any data synthesis. | 8 |
| Summary measures | 13 | State the principal summary measures (e.g., risk ratio, difference in means). | 9 |
| Synthesis of results | 14 | Describe the methods of handling data and combining results of studies, if done, including measures of consistency (e.g., $I^2$) for each meta-analysis. | 9 |

| Section/topic | # | Checklist item | Reported on page # |
| --- | --- | --- | --- |
| Risk of bias across studies | 15 | Specify any assessment of risk of bias that may affect the cumulative evidence (e.g., publication bias, selective reporting within studies). | 9 |
| Additional analyses | 16 | Describe methods of additional analyses (e.g., sensitivity or subgroup analyses, meta-regression), if done, indicating which were pre-specified. | 9 |
| **RESULTS** |  |  |  |
| Study selection | 17 | Give numbers of studies screened, assessed for eligibility, and included in the review, with reasons for exclusions at each stage, ideally with a flow diagram. | 10 |
| Study characteristics | 18 | For each study, present characteristics for which data were extracted (e.g., study size, PICOS, follow-up period) and provide the citations. | Table 1 |
| Risk of bias within studies | 19 | Present data on risk of bias of each study and, if available, any outcome level assessment (see item 12). | 11 |
| Results of individual studies | 20 | For all outcomes considered (benefits or harms), present, for each study: (a) simple summary data for each intervention group (b) effect estimates and confidence intervals, ideally with a forest plot. | Figures 2-4 |
| Synthesis of results | 21 | Present results of each meta-analysis done, including confidence intervals and measures of consistency. | Figures and Suppl. Figures 6-8 |
| Risk of bias across studies | 22 | Present results of any assessment of risk of bias across studies (see Item 15). | Table 2 |
| Additional analysis | 23 | Give results of additional analyses, if done (e.g., sensitivity or subgroup analyses, meta-regression [see Item 16]). | Figure 5 |
| **DISCUSSION** |  |  |  |

|  |  |  |  |
| --- | --- | --- | --- |
| Summary of evidence | 24 | Summarize the main findings including the strength of evidence for each main outcome; consider their relevance to key groups (e.g., healthcare providers, users, and policy makers). | 14 |
| Limitations | 25 | Discuss limitations at study and outcome level (e.g., risk of bias), and at review-level (e.g., incomplete retrieval of identified research, reporting bias). | 16 |
| Conclusions | 26 | Provide a general interpretation of the results in the context of other evidence, and implications for future research. | 17 |
| **FUNDING** |  |  |  |
| Funding | 27 | Describe sources of funding for the systematic review and other support (e.g., supply of data); role of funders for the systematic review. | Title Page |

From: Moher D, Liberati A, Tetzlaff J, Altman DG, The PRISMA Group (2009). Preferred Reporting Items for Systematic Reviews and Meta-Analyses: The PRISMA Statement. PLoS Med 6(6): e1000097. doi:10.1371/journal.pmed1000097
For more information, visit: www.prisma-statement.org.

**Supplementary Table 2:**
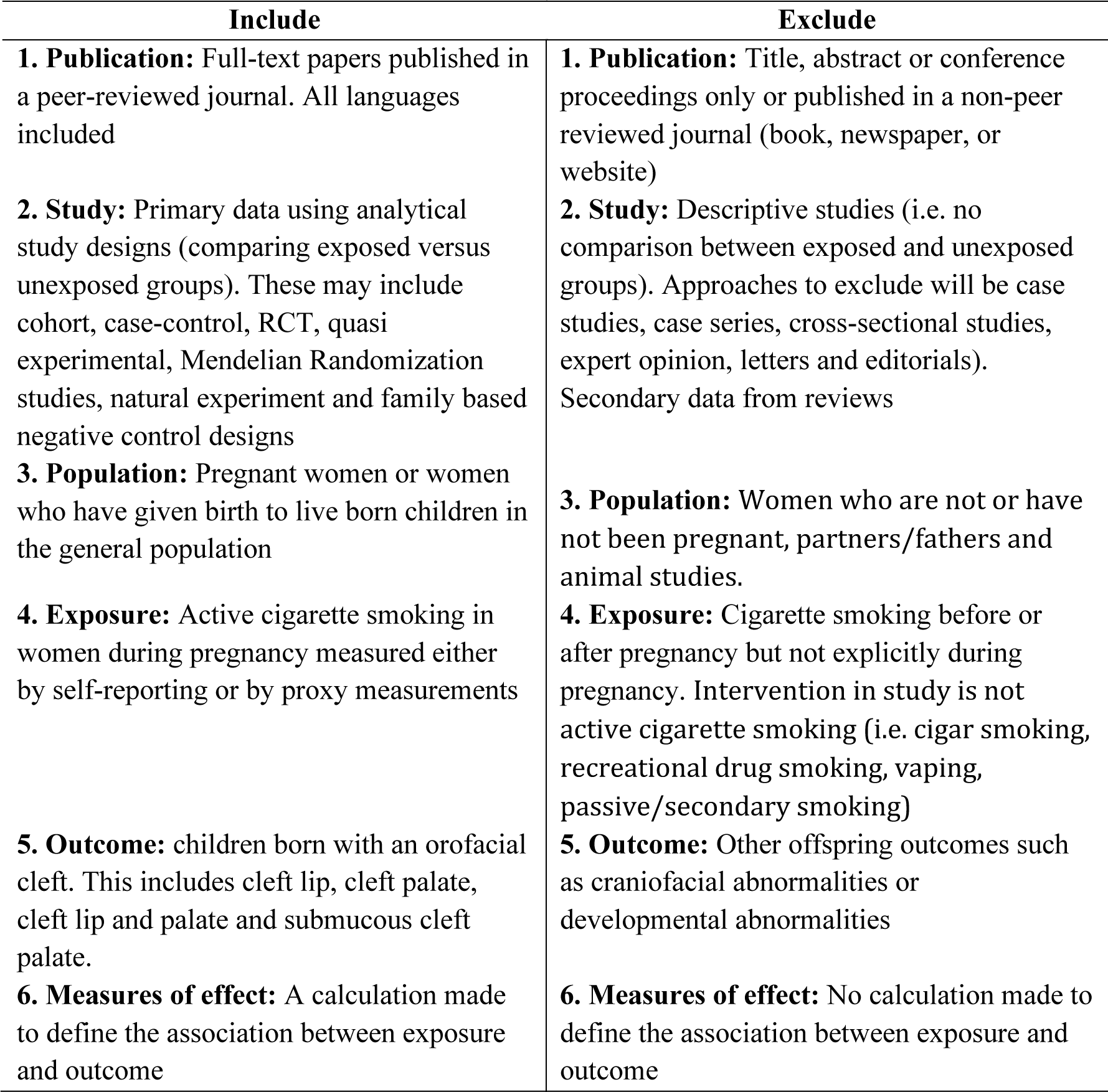
Criteria for including or excluding papers

**Supplementary Table 3:**
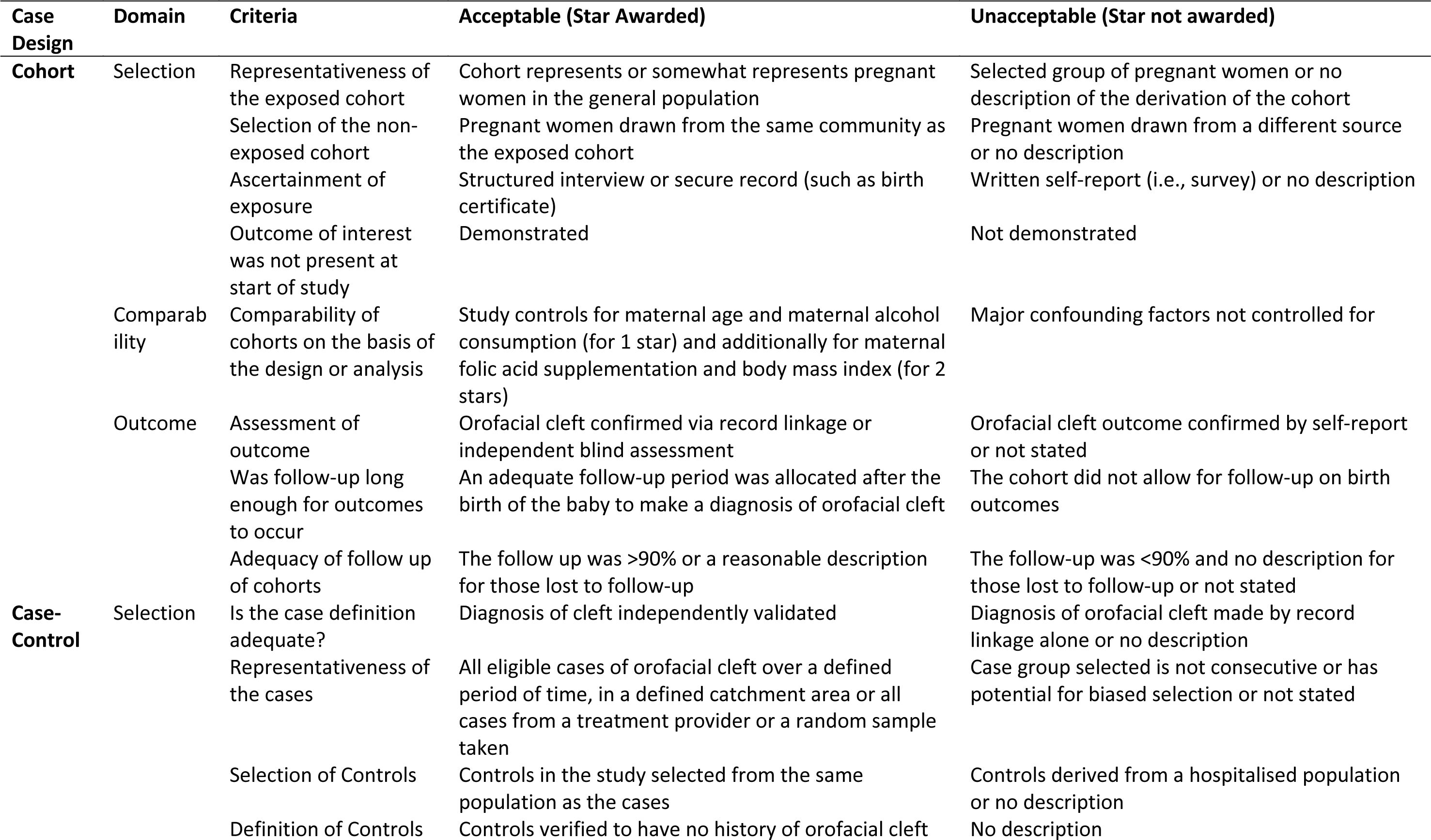
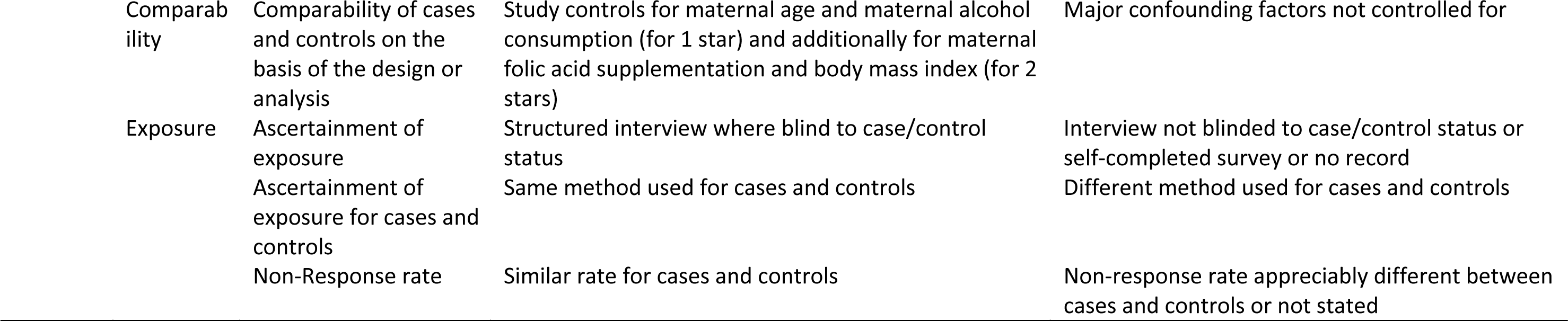
The Assignment of Stars for Study Quality using the Newcastle Ottawa Scale

**Supplementary Table 4:**
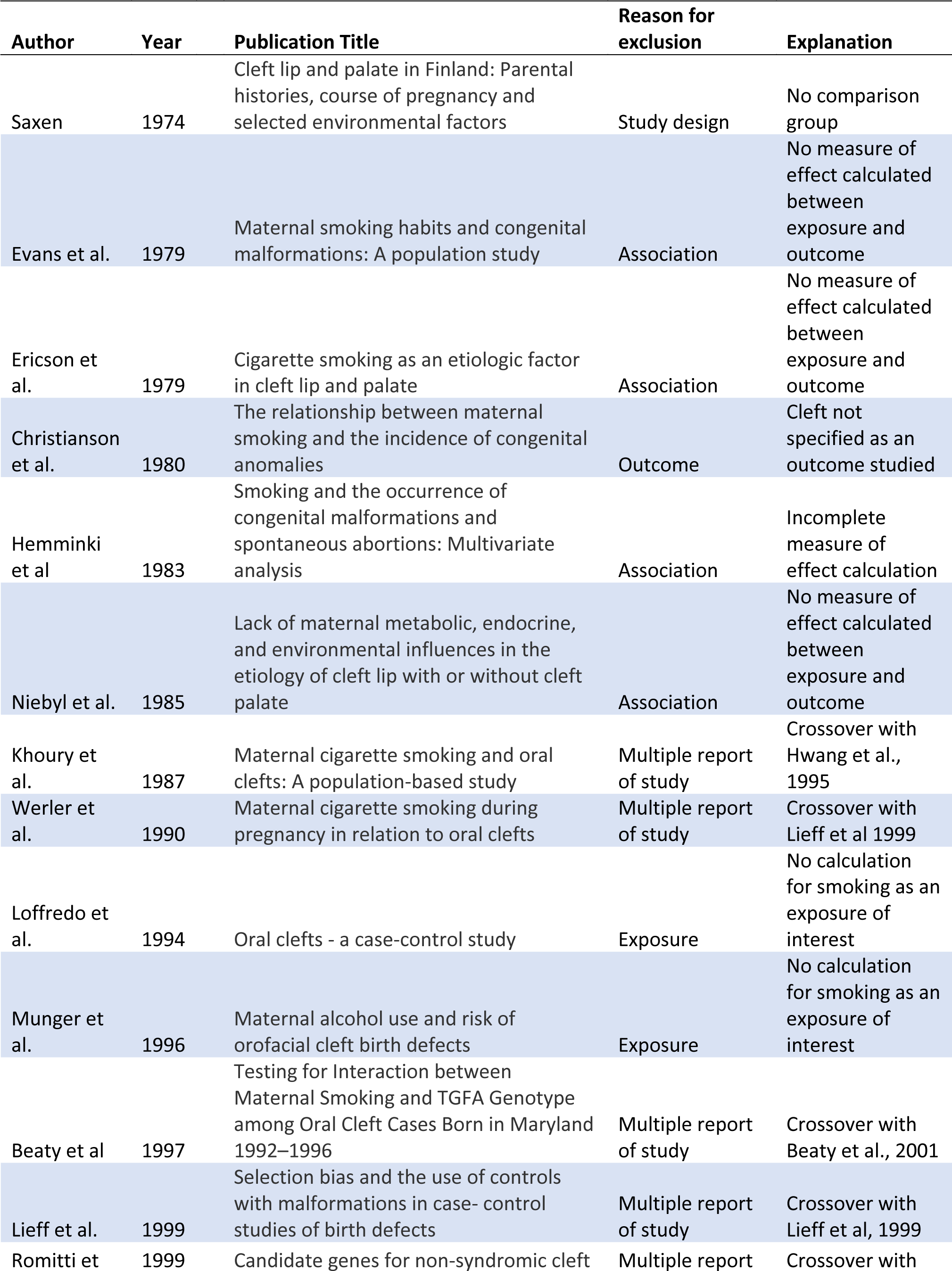
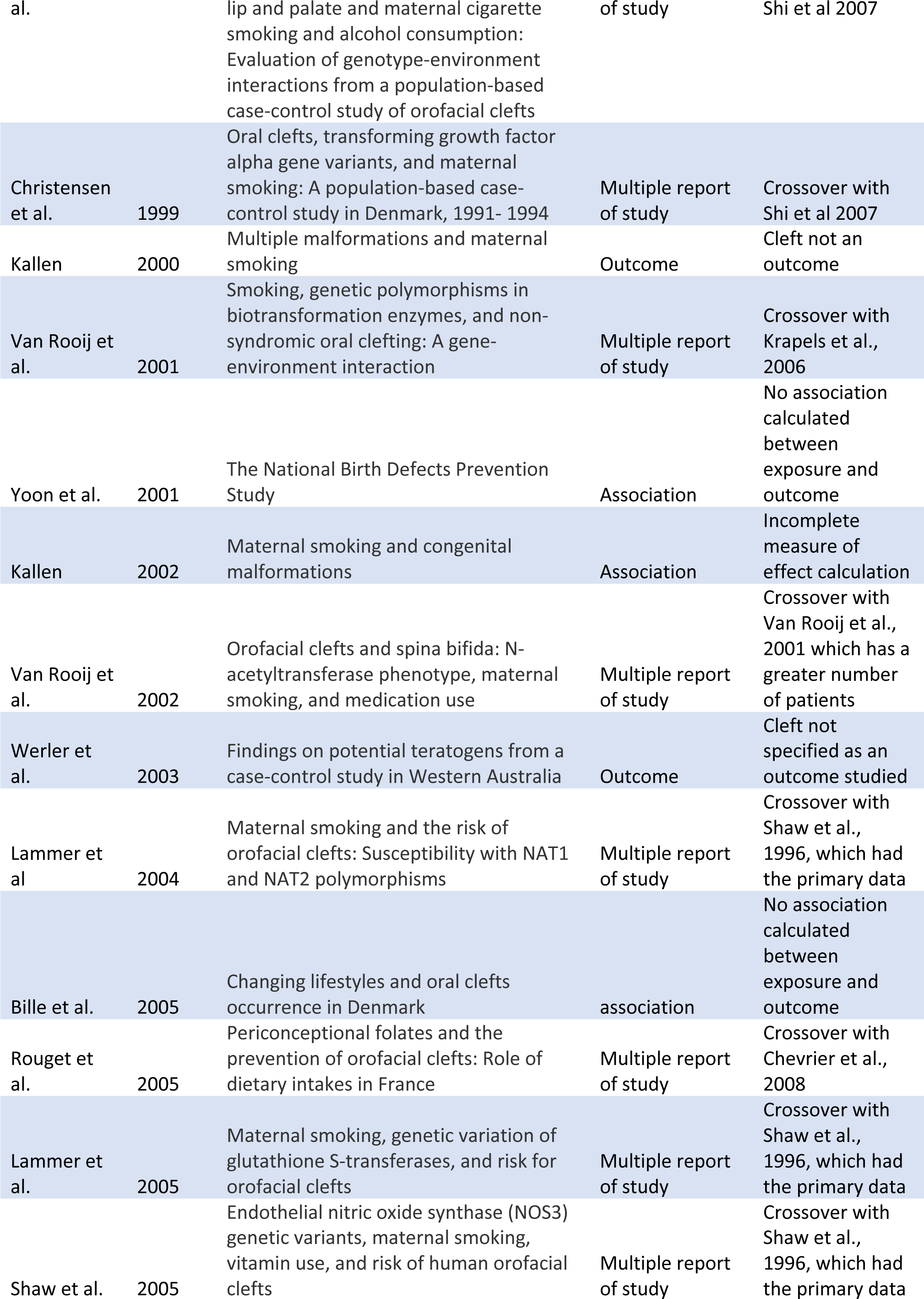
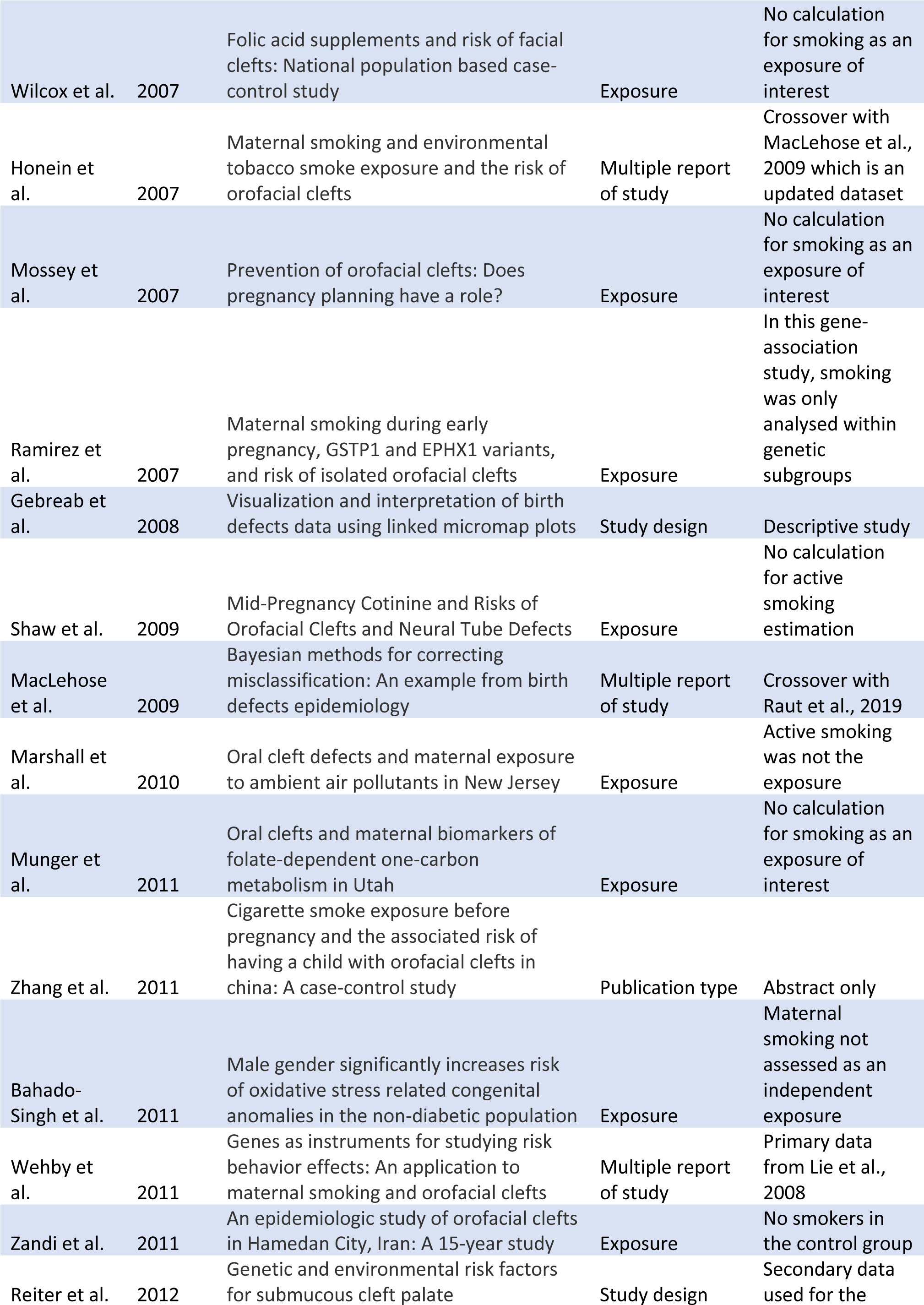
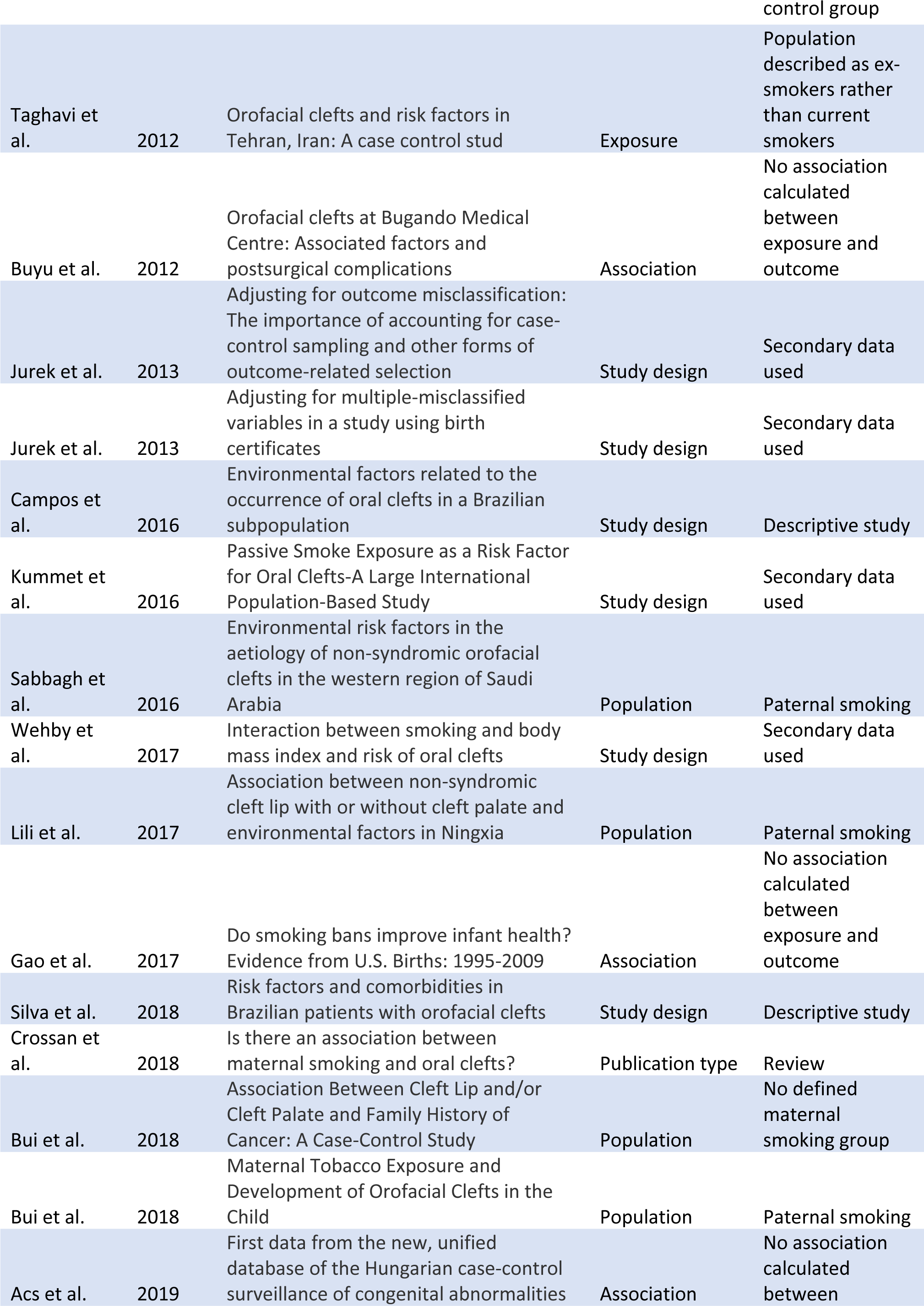
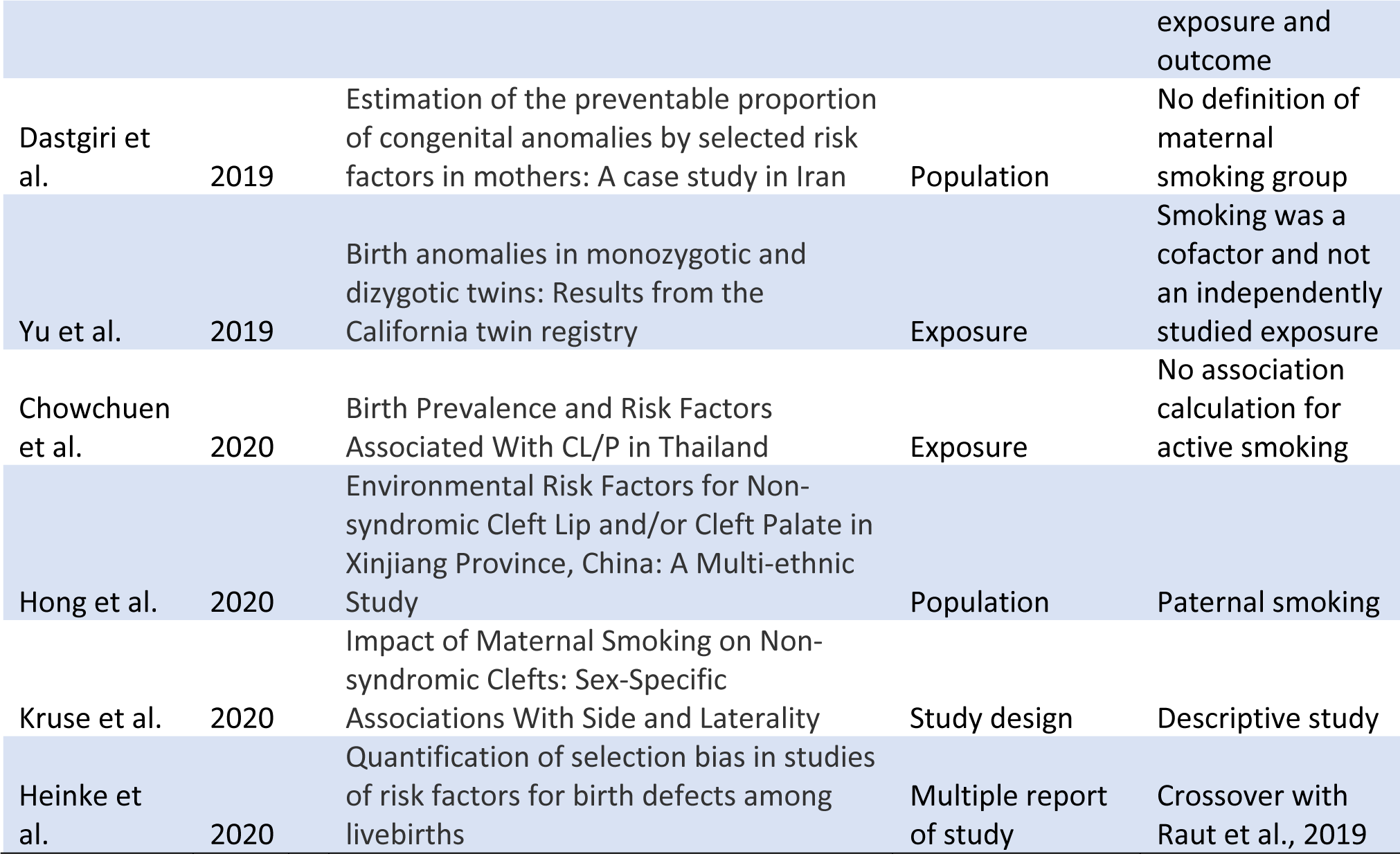
Articles Excluded at the Full Text Screening Stage and Reasons for Exclusion

**Supplementary Table 5:** Confounding Factors Adjusted For in All Included Studies

| Author | Year | Maternal age | Alcohol | Folic Acid | Obesity |
| --- | --- | --- | --- | --- | --- |
| Shiono et al. | 1986 | ◆ | ◆ |  |  |
| Khoury et al. | 1989 | ◆ | ◆ |  |  |
| Malloy et al. | 1989 | ◆ |  |  |  |
| Van Den Eeden et al. | 1990 | ◆ | ◆ |  |  |
| McDonald et al. | 1992 | ◆ | ◆ |  |  |
| Hwang et al. | 1995 | ◆ |  |  |  |
| Shaw et al. | 1996 | ◆ | ◆ | ◆ |  |
| Kallen | 1997 | ◆ |  |  |  |
| Lieff et al. | 1999 | ◆ |  |  |  |
| Lorente et al. | 2000 | ◆ | ◆ |  |  |
| Chung et al. | 2000 | ◆ | ◆ |  |  |
| Beaty et al. | 2001 | ◆ | ◆ | ◆ |  |
| Woods et al. | 2001 | ◆ |  |  |  |
| Wyszynski and Wu | 2002 | ◆ | ◆ |  |  |
| DeRoo et al. | 2003 | ◆ |  |  |  |
| Little et al. | 2004 |  | ◆ | ◆ |  |
| Meyer et al. | 2004 | ◆ |  |  |  |
| Krapels et al. | 2006 |  | ◆ | ◆ |  |
| Shi et al. A | 2007 |  | ◆ | ◆ |  |
| Shi et al. B | 2007 | ◆ | ◆ |  |  |
| Bille et al. | 2007 | ◆ | ◆ | ◆ |  |
| Grewal et al. | 2008 | ◆ | ◆ | ◆ | ◆ |
| Lie et al. | 2008 | ◆ | ◆ | ◆ |  |
| Chevrier et al. | 2008 | ◆ | ◆ | ◆ |  |
| Leite and Koifman | 2009 | ◆ | ◆ |  |  |
| Lebby et al. | 2010 | ◆ | ◆ |  |  |
| Mirilas et al. | 2011 |  |  |  |  |
| Zhang et al. | 2011 | ◆ | ◆ | ◆ |  |
| Wehby et al. | 2011 |  | ◆ | ◆ | ◆ |
| Ibarra-Lopez et al. | 2013 |  |  | ◆ |  |
| Gunnerbeck et al. | 2014 | ◆ |  |  |  |
| Leite et al. | 2014 | ◆ |  |  | ◆ |
| Salihu et al. | 2014 | ◆ |  |  |  |
| Bezerra et al. | 2015 |  | ◆ |  |  |
| Hao et al. | 2015 | ◆ | ◆ |  | ◆ |
| Martelli et al. | 2015 |  |  |  |  |
| Figueiredo | 2015 |  | ◆ |  |  |
| Ebadifar et al. | 2016 | ◆ |  |  |  |
| Liu et al. | 2016 |  |  |  |  |
| Angulo-Castro et al. | 2017 |  | ◆ | ◆ |  |
| Xu et al. | 2018 | ◆ | ◆ | ◆ |  |
| Raut et al. | 2019 | ◆ | ◆ | ◆ | ◆ |
| Sato et al. | 2020 | ◆ | ◆ | ◆ | ◆ |
| Acs et al. | 2020 | ◆ | ◆ |  |  |
| Regina et al | 2020 |  | ◆ | ◆ | ◆ |
| Ausländer et al. | 2020 | ◆ | ◆ | ◆ | ◆ |

